# Patterns of SARS-CoV-2 Aerosol Spread in Typical Classrooms

**DOI:** 10.1101/2021.04.26.21256116

**Authors:** Gerhard K. Rencken, Emma K. Rutherford, Nikhilesh Ghanta, John Kongoletos, Leon Glicksman

## Abstract

Although current industry guidelines to control the spread of SARS-CoV-2 (COVID-19) have adopted a six-foot (∼1.8m) spacing between individuals indoors, recent evidence suggests that longer range spread is also responsible for infections in public spaces. The vehicle for long-range spread is smaller droplets or particles, termed bio-aerosols, or aerosols for short, which have a large surface area to volume ratio such that aerodynamic drag is much larger than gravity forces. The aerosols remain suspended in air for extended time periods and they essentially move with air currents. Prediction of the danger to occupants in a closed room when exposed to an infected individual requires knowledge of the period of exposure and the concentration level of aerosols in the breathing zone of an occupant. To obtain an estimate of the concentration level, a common assumption is well-mixed conditions within an interior space. This is obtained from a mass balance between the level of aerosol produced by an infected individual along with the airflow rate into and out of the entire space. In this work, we use computational fluid dynamics, verified by experimental results, to explore the aerosol concentration distribution in a typical classroom for several common conditions and compare these results to the well-mixed assumption. We use a tracer gas to simulate the flow and dispersion of the aerosol-air mixture. The two ventilation systems examined, ceiling diffusers and open windows, yield average concentrations at occupant breathing level 50% greater than the well mixed case, and some scenarios yield concentrations that are 150% greater than the well mixed concentration at specific breathing-level locations. Of particular concern are two conditions: horizontal air flow from an open window in line with a row of seating and, second, an infected individual seated near a sealed cold window. For the former, conditions are improved if a baffle is placed inside the open window to direct the air toward the floor, creating a condition similar to displacement ventilation. In the latter, the cold air flowing down along the cold window recirculates aerosols back into the breathing zone. Adding window covers or a portable heater below the window surface will moderate this condition.

## Introduction

SARS-CoV-2 (COVID-19) has fundamentally altered many aspects of our daily lives. Despite this impact and the enormity of the research that has gone into prevention strategies, many of the industry guidelines currently in use assume a well-mixed air model [1, 2], which holds that the aerosol concentration is uniform throughout a room volume at a specific moment in time. This assumption has inherent flaws regarding both personal protection and energy consumption. Building on government guidelines and published literature, six-foot (∼1.8m) spacing between individuals and the wearing of face masks are now generally accepted practices for reducing transmission rates [3, 4, 5]. However, recent evidence has suggested that longer range spread is responsible for infections in public spaces such as restaurants [6], public transit [7], hotels [8], hospitals [9], multistory buildings [10, 11], and cruise ships [12, 13, 14].

Whereas earlier studies suggested that SARS-CoV-2 spread was due to large drops dispersed within a rough ballistic trajectory of 3 to 6 feet or by surface contamination [15], more recent work has highlighted the vehicle for long-range spread as smaller droplets or particles, termed bio-aerosols, or aerosols for short, which are assumed to remain suspended in air for extended time periods [16, 17, 18, 19]. It is even possible that the aerosols are more infectious than larger particles [20]. The upper limit for the size of these bio-aerosols is 5 µm [21, 22, 15, 23], with a possibility of particles less than 100 to 200 µm becoming aerosols through evaporation [24]. As these aerosols can settle with time, an assumption of the settling velocity can be found from the Stokes equation for the drag between particles and surrounding air [25], assuming a spherical shape and density of liquid water. The Stokes equation suggests that the relative velocity between the particle and the surrounding air is less than 10^−3^ m/s, or rather, that the bio-aerosols follow the trajectory of the room air currents. Such concerns are not new [26, 27] and serve to raise questions about understanding the efficacy of ventilation systems [28, 29, 16, 30, 31].

In order to establish proper safety guidelines for interior spaces, it is important to predict the danger to occupants when exposed to an infected individual. This requires a knowledge of the period of exposure and the concentration level of aerosols in the breathing zone of an occupant. Some investigators combine this knowledge with the Wells-Riley equation [32, 33, 34] to predict the conditional probability of infection [35, 32, 4, 36]. To obtain an estimate of the concentration level, a common assumption is well-mixed conditions within an interior space [37, 33]. Given the level of aerosol produced by an infected individual along with the heating, ventilation and air conditioning (HVAC) flow rate and room geometry, a simple mass balance will yield the time resolved concentration level within the space. While the well-mixed values can lend valuable information regarding the air change rate necessary to ventilate a space, we have reason to believe that room specific conditions such as ventilation type and the locations of individuals can have large effects on the distribution of aerosol in the room.

There are several documented cases of infections of individuals seated far from the original source. The new infections are postulated to be caused by aerosol concentrations around the infected individuals well above the average for the entire room [7, 11]. Li documents the probable aerosol transmission of CoV-2 across the width of a crowded restaurant in Guangzhou due to the draft from a wall air conditioner [6, 38]. Kwon [39] presents evidence of the transmission of SARS-CoV2 across the width of a South Korean restaurant where an individual was infected from approximately 6.5 m away. Subsequent measurements of the air flow in the restaurant revealed a horizontal air stream originating from a ceiling diffuser, moving down a wall, and then being deflected by a table to form a horizontal flow with a velocity of about 1 m/s. The air stream passed close to an infected customer seated near the wall and then across the room to a second customer, who became infected. Due to this evidence of location dependent infection, we analyze both the well-mixed concentrations as well as the situation-specific hotspots of high aerosol concentration.

The challenge arises to characterize aerosol dispersion patterns for several typical interior spaces under different operating conditions. When is the well-mixed assumption valid? Under what conditions are occupants potentially exposed to unsafe high concentration and where are the unsafe locations? This, in turn, requires a generalized understanding of air flow patterns which is lacking in the current literature.

In the past, detailed studies of airflow behavior in interior spaces have been carried out, both experimentally [40] and computationally [41], to characterize natural ventilation patterns. People at rest generate about 75 watts, heating the surrounding air and creating rising plumes of warm air [42]. These plumes have a typical velocity on the order of 0.1 to 0.3 m/s. As they rise, they entrain cooler surrounding air and continue up until they encounter a layer of air at the same temperature. At this layer, the plumes will dissipate and mix with the surrounding air [43, 44]. Walls and floors heated by solar gains can also warm nearby air to create positively buoyant flows while cold windows create downflowing cold films with negative buoyancy. These buoyant flows, created by heat fluxes into and out of the room, interact with airflow from windows, vents, HVAC supply ducts, diffusers, and other heated and cooled objects in the room.

HVAC systems are designed to limit the internal air velocity around humans to reduce discomfort due to draft. Discomfort is observed generally when the air velocity reaches the range of 0.3 m/s and above [42]. Therefore, the thermal plumes and mechanical air flows have velocities that are of the same order of magnitude and together dictate the overall air flow pattern in interior spaces. The only exception is air flow from windows under high wind conditions and possibly flow created by ceiling fans that can dominate the internal flow behavior. Results from natural ventilation studies have revealed a complex pattern of air flow in buildings. In some cases, a thick heated layer of air is present below the ceiling. In others, there are downward flowing streamlines of cooler air or a series of large swirling vortices [41].

There is a dearth of systematic studies of aerosol distribution in common interior spaces, other than a limited number for spaces such as small offices [45], hospitals [46], and aircraft cabins [47]. Two recent studies have addressed safe ventilation in classrooms [48] and outdoor dining [49]. In the former, the use of ventilation from an open window above the breathing level in a typical New York classroom creates a safer condition. To broaden our understanding, we have carried out a detailed study of aerosol concentration patterns in a common space: a classroom with and without an operable exterior window supplemented by a study of streamline flow patterns in a conference room. Our goal is to find the spatial concentration distribution produced by an infected individual under several different operating and seasonal conditions. Attention will be focused on the aerosol concentration other classroom occupants breathe. We use a tracer gas to simulate the flow and dispersion of the aerosol-air mixture. The simulations assume steady state conditions. At air change rates of 4 to 6 air changes per hour, conditions are close to steady-state within one half of a normal hour-long session. We will assume that the tracer concentration levels will scale linearly with the source concentration and the tracer/aerosol moves and diffuses with the surrounding gas flow patterns. To answer the question regarding the accuracy of the well mixed assumption, the local concentration at the breathing level of the occupants will be scaled to the steady state well mixed value.

## Computational Fluid Dynamics

### Governing Equations

To computationally estimate the airflow within the space, computational fluid dynamics (CFD) was used. ANSYS Fluent software was used to simulate both airflow patterns and aerosol concentration in the conference room and classroom defined below. A steady state solver, the Reynolds Average Navier Stokes (RANS) with pseudo-transient effect was used to solve for the airflow pattern [50].

A single-phase flow with mass and momentum conservation was selected, coupled with the k-epsilon turbulence model with enhanced near-wall function to solve the effects of turbulence. The Species Transport model was used to solve for the differing mass concentration of different gaseous species in the space, and the Surface to Surface Radiation model with diffuse surfaces was used to calculate the radiation heat transfer component.

The mass conservation equation is given by:

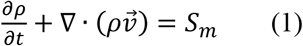

The right-hand term is the mass added to the continuous phase from the dispersed second phase (such as user-defined mass sources).

The solver solves the energy equation in the form below:

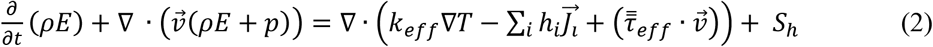

Where *k*_*eff*_ is the effective conductivity, 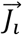 is the diffusion flux of species i, *Y*_*i*_ is the mass fraction of species i, and

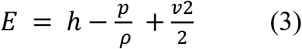

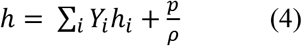

The momentum conservation equation is solved in the following form:

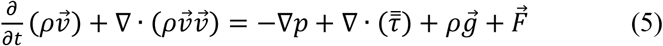

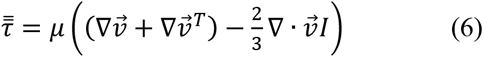

Where: *p* is the static pressure, 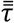 the stress tensor, 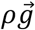 the gravitational body force, 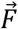 the external body force, *μ* the molecular viscosity, *I* the unit tensor, and 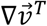 the effect of volume dilation.

To predict the aerosol concentration at different locations in the classroom, we make use of the species transport model, that predicts the local mass fraction of each species *Y*_*i*_ by obtaining the solution of a convection-diffusion equation for the i^th^ species. This equation is given, in the general form, as:

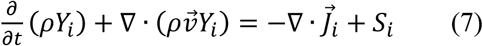

Where *S*_*i*_ is the rate of generation of a species at user-defined sources (diffuser inlets, windows and mouths).

This equation is solved n-1 times, where n is the total number of species. Since the mass fractions sum to unity, the final species is calculated by finding the remaining mass fraction that has not yet been solved for. To reduce numerical error, the species with the highest mass fraction is selected as the last species (in our case nitrogen).

In turbulent conditions the mass diffusion, 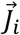, is computed as follows:

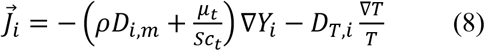

Where *Sc*_*t*_ is the turbulent Schmidt number (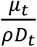 where *μ*_*t*_ is the turbulent viscosity and *D*_*t*_ is the turbulent diffusivity).

To model the effects of turbulence in the simulation, we utilize the standard k-epsilon model. This two-equation model allows the solver to compute for both a turbulent length as well as time scale. The equations provide a reasonably robust and economic accuracy for a variety of turbulent flows.

The Surface-to-Surface (S2S) model was used to account for the radiative heat transfer between surfaces in the room. This model takes into consideration the size, separation distance, and orientation of surfaces, through a calculated view factor, while ignoring any effects of absorption, emission or scattering. All surfaces are assumed to be diffuse emitters and reflectors by use of radiosity.

### Geometry

Our base classroom geometry was abstracted from a classroom at MIT which is 12 meters long, 6 meters wide, and 3.5 meters tall, as shown in Figure 1. Under the assumption that precautions for safety during COVID-19 would be considered when setting up a classroom, we placed ten students into two rows, all at least six feet apart (∼1.8m). In some simulations, we considered a large single glazed window behind the back row of students, in others, open windows on one end wall.

**Figure 1:**
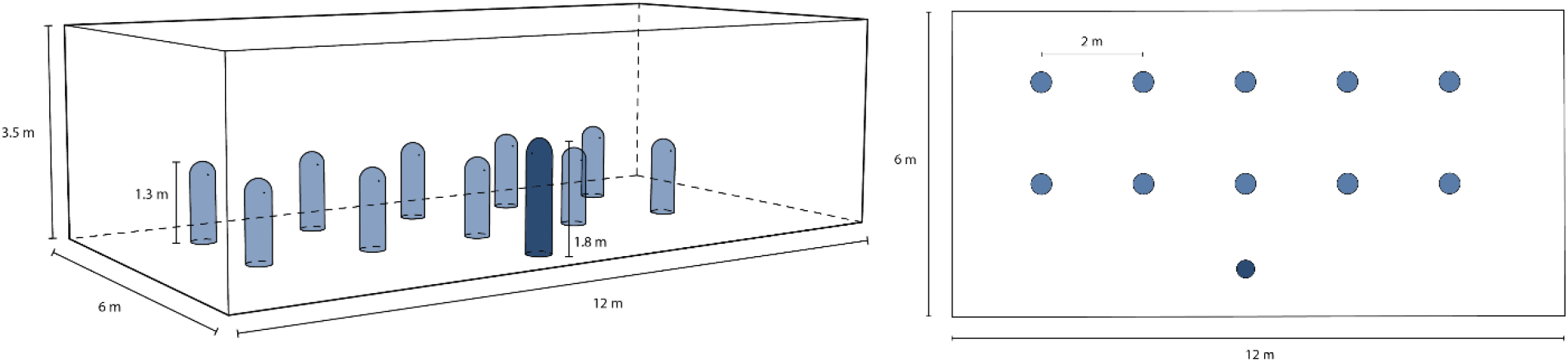
Classroom geometry for simulations. People are modelled by 1.3 m tall cylinders with hemispheres on top. They are placed in two rows so that all students are six feet apart.

We used a simplified human geometry of a cylinder with a half sphere on top due to this forms accuracy in modeling plumes. Students were modeled to be seated at a height of 1.3 meters with a standing teacher at the front of the classroom at a height of 1.8 meters. The students had a surface area of 1.7 m^2^ and the teacher a surface area of 1.9 m^2^. Each person has a circular mouth of variable area, based on the exhalation velocity.

### Boundary Conditions

All walls, the floor, and the ceiling were assumed to be adiabatic unless otherwise stated. The humans were given a heat transfer rate of 44 Watts/m^2^ (convection plus radiation) which provides 75 Watts from each student, a typical heat generation rate for a resting person, and 84 Watts from the teacher, corresponding to a standing, more active person.

Each mouth was simulated as as a constant velocity source of either 0.35 m/s or 1.0 m/s normal to the surface, resulting in a mass flow rate of 0.42 m^3^/hour, which corresponds to an average human exhalation rate. The temperature of the exhaled air is set at human body temperature of 310 K. To track the mixture of aerosols and air exhaled by an “infected” individual, we chose to use carbon monoxide (CO) as a tracer gas due to its similar density and viscosity to air. The infected individual’s exhaled air was 100% CO and the uninfected individuals’ exhaled air was 16% oxygen (O_2_), 4% carbon dioxide (CO_2_), and 80% nitrogen (N_2_). This enabled us to visualize the possible spread of virus-infected aerosols throughout the room and measure the concentrations at various locations to compare between models.

In this paper, we look at various combinations of ventilation methods and room conditions, including HVAC through ceiling diffusers, opening windows and doors, and non-adiabatic closed windows with temperature differences across them. It is assumed that all supply air does not contain any infected aerosols. In some simulations, a negative or positive heat flux is introduced to the rear wall of the classroom to model a single-glazed window during either hot or cold seasonal conditions. In natural ventilation simulations, windows are introduced on the left (-X direction) wall, which are modeled by rectangular cutouts on the left wall.

### Experimental Confirmation

To verify that the models and boundary conditions used in the CFD simulations resembled real world results, a case of a single person in a room was considered. In this case, our sitting human model of a cylinder with a half sphere on top is positioned alone in a room. The results were compared to experimental measurements for the identical case [51] and are illustrated in Figure 2. The close agreement between the plume velocity at different distances above the head and the vertical temperature distribution in the room help to establish confidence in the computational results.

**Figure 2:**
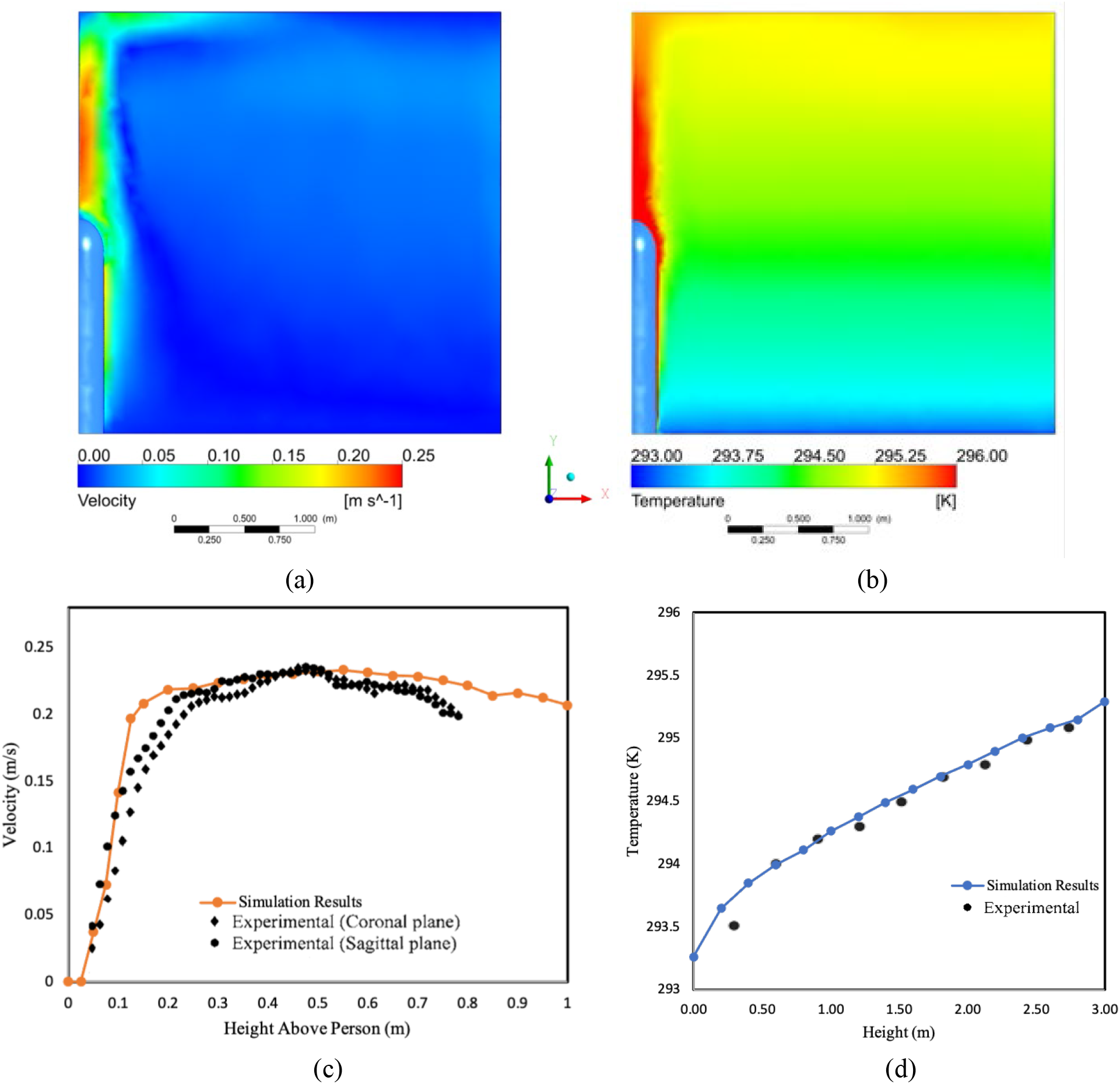
Experimental Confirmation of CFD Accuracy through comparison of plume rise velocity and temperature data from an experiment with single human subject with the results of our simulation modelling the same situation. Simulation results of (a) velocity distribution and (b) temperature distribution. Graphs of (c) plume velocity vs height compared with experimental data[42] above the head of a person and (d) temperature versus the height above the floor in the room, measured a meter away from the person.

A related study of air flow patterns in a conference room near MIT provides further confirmation of the accuracy of the CFD model used. The room studied in this related experiment had seating for six around a central table, a chilled beam conditioning system, a larger double-glazed window, and exhaust through the doorway. Both the conference room [52] and classroom studies were modeled in Ansys Fluent with similar boundary conditions to the ones we used. Experimental measurements of the conference room were able to be taken to gain confidence in the results. Near the doorway the experimental velocity varied by as much as 0.5 m/s from floor to ceiling. The simulated velocity values at this location agreed with the experimental ones within six percent on average, while the simulated vertical temperature profile agreed within 0.5 °K with measurements.

## Results

### Effects of Exhale Speed on Direction of Expelled Aerosols and Importance of Masks

When researching the spread of virus containing aerosols in an indoor environment, the importance of human plumes is often overlooked. We argue, however, that warm air plumes drastically affect the movement of air in rooms with stationary people. Exhaled air is immediately affected by the warm air plume rising around a human, and the speed of the exhalation determines whether or not the aerosols within a breath rise with the plume towards the ceiling or escape the plume and follow other air currents in the room. We assume in our simulations that air leaves the mouth horizontally at human body temperature with a mass flow rate comparable to the exhalation speed of a resting person. The horizontal velocity of the infected air escaping from the mouth can vary greatly depending on the presence and fit of masks.

To examine the effects a mask might have in diverting infected air away from other individuals, a simulation involving discrete water droplets of varying initial velocities was run. In this setup, there is a single individual in a room, without any flow generated by the HVAC system. To reach steady state, the side walls have a negative heat flux such that the total heat generated by the human is removed by the walls. The walls also act as a pressure boundary condition at gauge pressure, allowing air to pass out of the room. Discrete water droplets with a 1 μm diameter were injected into the room from the mouth so that they could be carried by exhaled air of varying speeds. Figure 3 shows three paths of water droplets with exhale speeds of 0.35 m/s, 1.0 m/s, and 2.0 m/s, colored by velocity as they leave the mouth and rise.

**Figure 3:**
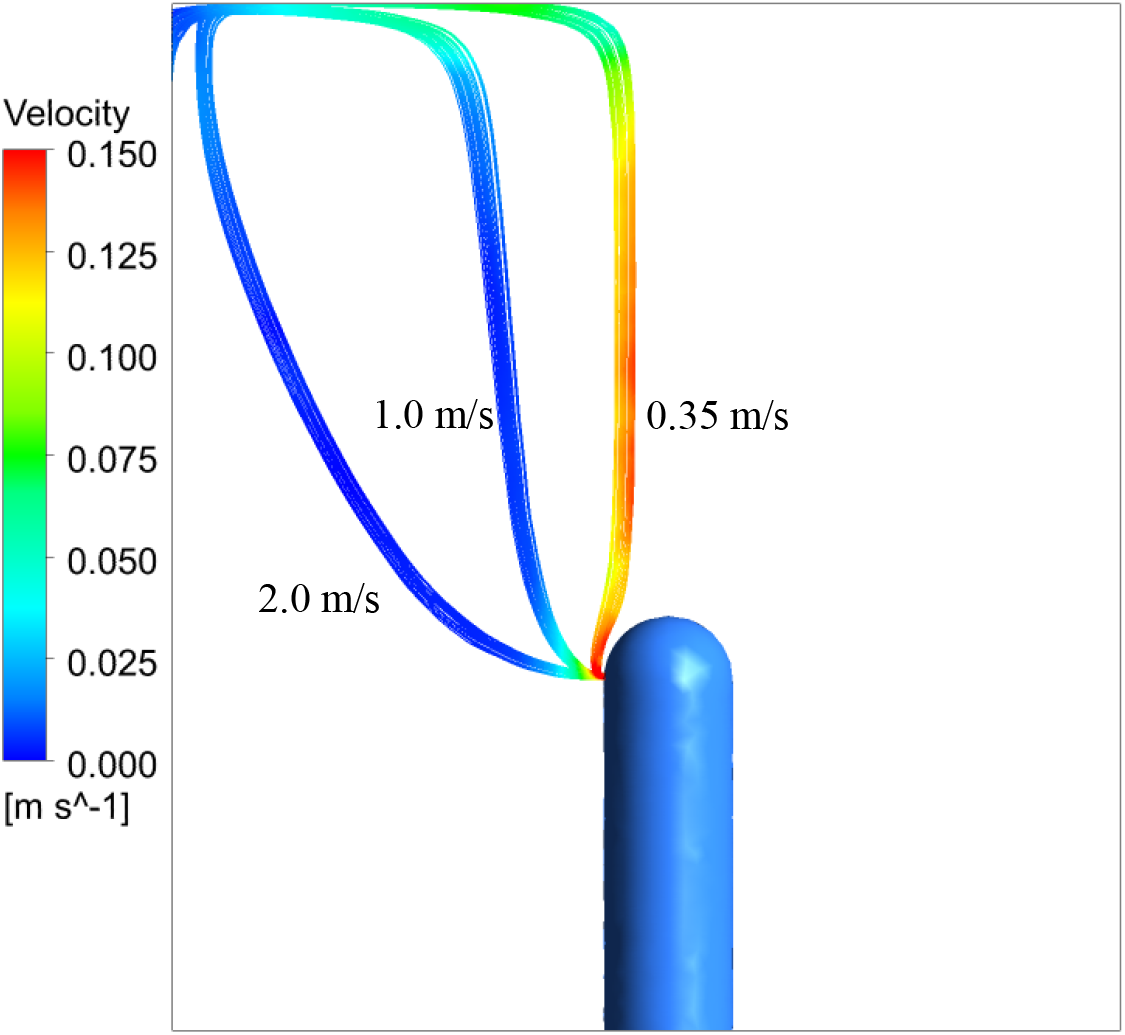
Trajectories of water droplets exhaled at various speeds. The leftmost stream corresponds to an exhale velocity of 2 m/s, which is a usual value for the initial velocity of a human breath. The middle stream was exhaled at 1 m/s, which is a speed in between a masked and unmasked case. The rightmost stream was exhaled at 0.35 m/s, corresponding to very little horizontal velocity due to a well-fitting mask being worn. The color of the streams indicate their velocity.

We found that only air leaving the mouth area at a very low speed, comparable to the speed of air being exhaled through or around a mask, rises with the human plume. All greater exhalation speeds have a high enough velocity to escape the plume. The exhaled air is hotter than the surrounding air, so it will rise, although at a lower velocity than the particles in the plume. This is seen in Figure 3, with the rising velocity of the 2 m/s and 1 m/s particles being approximately 0.01 m/s and the rising velocity of particles in the plume being approximately 0.125 m/s. Therefore, the aerosols that escape the plume have a high chance of interacting with other air circulation in the room and spending more time at the breathing level or below rather than rising to the ceiling, creating a more infectious environment. These results further emphasize the importance of wearing a mask in indoor environments.

The CFD model for the conference room [52] illustrates this same phenomenon. In this simulation, exhaust from the individual mouths was omitted. Figure 4(a) shows streamlines of air originating in the thermal plumes right above the head of the two individuals sitting on the left of the table. Figure 4(b) depicts a streamline starting 5 cm away from the seated individual’s head. The distinction between these flow paths despite relatively close starting locations shows the importance of the plume in redirecting air. It also shows how thin the thermal boundary layer producing the plume is to the body since air only 5 cm away did not experience the buoyant effects. This supports the idea that a low exhale velocity is needed for infected breath to be captured by the human plume. Therefore, aerosols emitted around a face mask that stay within the buoyant plume have a different trajectory than aerosols expelled with a horizontal velocity that takes them outside the human plume.

**Figure 4:**
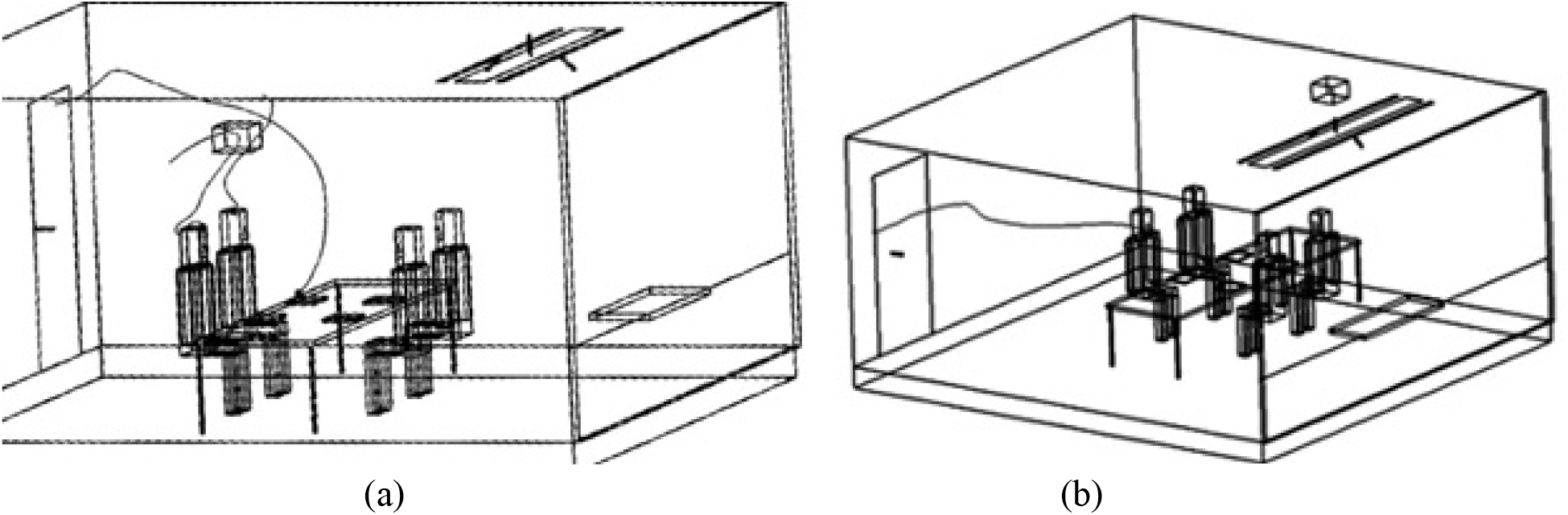
Streamlines originating (left) within the thermal plumes and from a computer, and (right) 5 cm from the face.

As a comparison to these thermal plume effects, we also ran a simulation of the ceiling diffusers running in a classroom with adiabatic walls and no people in the room. In this case, streamlines originating from the inlets show a fairly random pathway of air throughout the space, compared to the streamlines present in a classroom with people, where air is warmed and rises around the people and then falls again as it cools. The significance of the thermal plumes on the overall airflow pattern in the room is further supported by these results.

### Display of Results

Both qualitative and quantitative displays are used to illustrate the results. Methods to compare the simulations include volume renders and color-coded planes of temperature and CO mass fraction, as well as streamlines showing air paths from the infected individuals. To easily compare results between simulations, we will use a scale relative to the calculated well-mixed concentrations of the room with the given boundary conditions. For the Natural Ventilation Simulations, volume renderings are displayed with transparency representing twice the well-mixed concentration and red representing four times the well-mixed concentration. Plane views of the seated and standing breathing planes are colored with blue representing areas at or below the well mixed concentration and red representing four times the well-mixed concentration. For HVAC and chilled beam ventilation the transparent limit for volume renderings is set at well-mixed concentration, and the opaque scale at four and a half times well-mixed concentration. Any point that has a mass fraction value below the transparent bound, is shown as transparent, enabling a 3D visual. This allows us to visually distinguish areas in the room with elevated aerosol concentrations.

### Natural Ventilation Results

Although most modern buildings are equipped with HVAC systems, many older buildings and schools rely on opening windows and doors for ventilation. Furthermore, many classrooms with mediocre ventilation systems are opening windows in hopes of increasing airflow and reducing the spread of COVID-19. For these reasons, it is important to consider the various scenarios that may arise when windows are opened and to analyze possible risk factors. Multiple setups with different window and door conditions were considered.

In the following natural ventilation simulations, we assume an exit velocity of 1 m/s for air breathed out by the students, corresponding to a poorly fitted mask case. We placed two infected individuals in the room, one close to the windows in the back row, and one far from the windows in the front row, so that we can compare the differing situations of being seated near or far from the inlet windows.

For the natural ventilation simulations, the same volumetric flow rate across the windows and through the mouths of the students is maintained. The total area of window inlets is 0.46 m^2^ and the velocity of incoming air is 1 m/s. This yields an air change rate of 6.5 changes per hour and a well-mixed, steady state concentration of 0.00052 total mass fraction of CO. The different situations modeled are described in Table 1 below.

**Table 1:** Descriptions of simulation variations with operable windows

|  |  |
| --- | --- |
| Simulation A | Double Hung Windows Aligned with Rows of Students |
| Simulation B | Double Hung Windows Between Rows of Students |
| Simulation C | Baffles on Double Hung Windows Aligned with Rows of Students |
| Simulation D | Door Open and Bottom Windows as Inlets Between Rows of Students |
| Simulation E | Door Open and Top Windows as Inlets Between Rows of Students |

#### Simulation A: Double Hung Windows Aligned with Rows of Students

In Simulation A, the windows are double hung and in line with the two rows of students, with an air speed of 1 m/s across each, normal to the boundary. Based on the nature of double hung windows, we assume a constant inflow of air through the bottom pair of windows at 1 m/s and set the top windows to be pressure outlets. There is no other exhaust vent. This situation would be similar to placing a fan near the lower windows or a breezy day with an overhang blocking the wind approaching the upper windows and the door closed. The results are pictured in Figure 5, with infected individuals indicated with an arrow.

**Figure 5:**
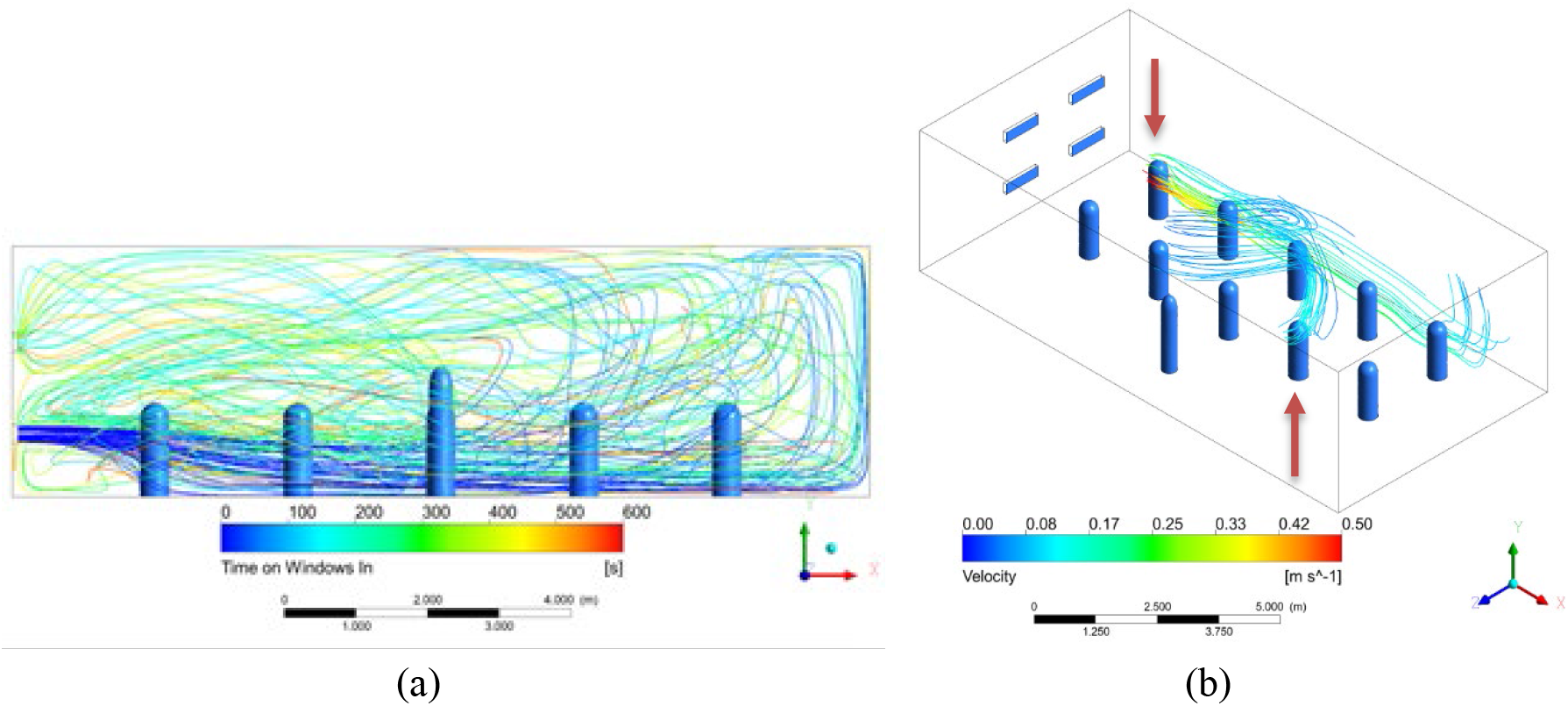
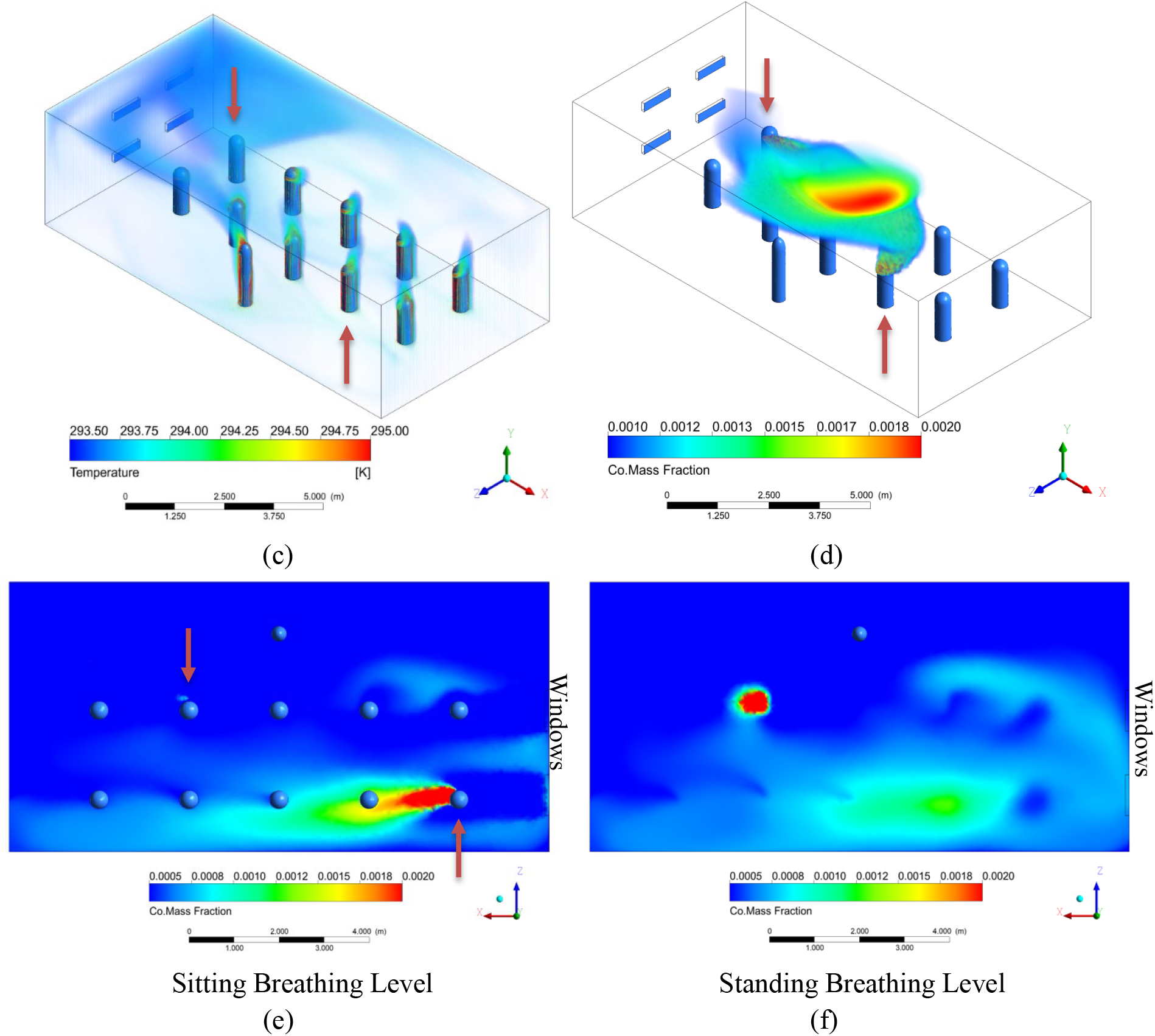
Results of Simulation A with double hung windows aligned with the rows of students. (a) Streamlines originating at bottom inlet windows from 0 to 600 seconds. (b) Streamlines originating around the heads of the two infected individuals from 0 to 60 seconds and colored according to velocity. (c) Volume rendering of temperature (K). (d) Volume rendering of CO mass fraction. The transparent limit is set at 2x well mixed, and the opaque scale at 4x well mixed. (e) Colored rendering of CO mass fraction right below mouth level of seated students. (f) Colored rendering of CO mass fraction right below mouth level of standing teacher. The lower limit is set at 1x well mixed, and the upper limit at 4x well mixed.

Our results in Figure 5(a) show a circular airflow pattern as cool air enters through the windows and sinks due to buoyancy effects while maintaining a horizontal velocity. As the air is warmed by the students and teacher, it rises and is pulled back toward the upper outlet window due to the positive pressure difference created by the incoming air. As seen in Figure 5(b) and Figure 5(e), this creates a dangerous situation when an infected student is seated next to the window, as infected aerosols are pushed directly into the next student by horizontal currents. Since the incoming air is cooler, the airstream drops as it moves toward the next student. If the open window was slightly higher, the concentration around the next student might be higher since the incoming air will blow rising aerosols from the infected individual’s plume and reach the next student at breathing level. The infected student farther away from the window does not create a hazardous environment for the person sitting next to them, and we mainly see the infected aerosols they breathed out rise to the ceiling.

#### Simulation B: Double Hung Windows Between Rows of Students

We hypothesized that this problem of horizontal breezes pushing infected air directly into other students could be mitigated by positioning the students so that they are not directly in line with the windows. We studied this case in Simulation B, which has the same setup as Simulation A, however, the windows were moved to be in the gaps between the two rows of students.

The resulting streamlines are very similar to in Simulation A, exhibiting a strong circular pattern of circulation in the XY plane, with the cool incoming air less disturbed by immediate interaction with the human plumes. In contrast to Simulation A, however, the plumes coming off the students in Simulation B are being largely pulled back towards the windows, see Figure 6(b), whereas the plumes were being largely pushed directly toward the next student in Simulation A. In this case, the students sitting next to the infected individuals are more protected than those in Simulation A, as seen by comparing Figure 5(e) to Figure 6(e). This suggests that seating students away from strong horizontal air currents is preferable.

**Figure 6:**
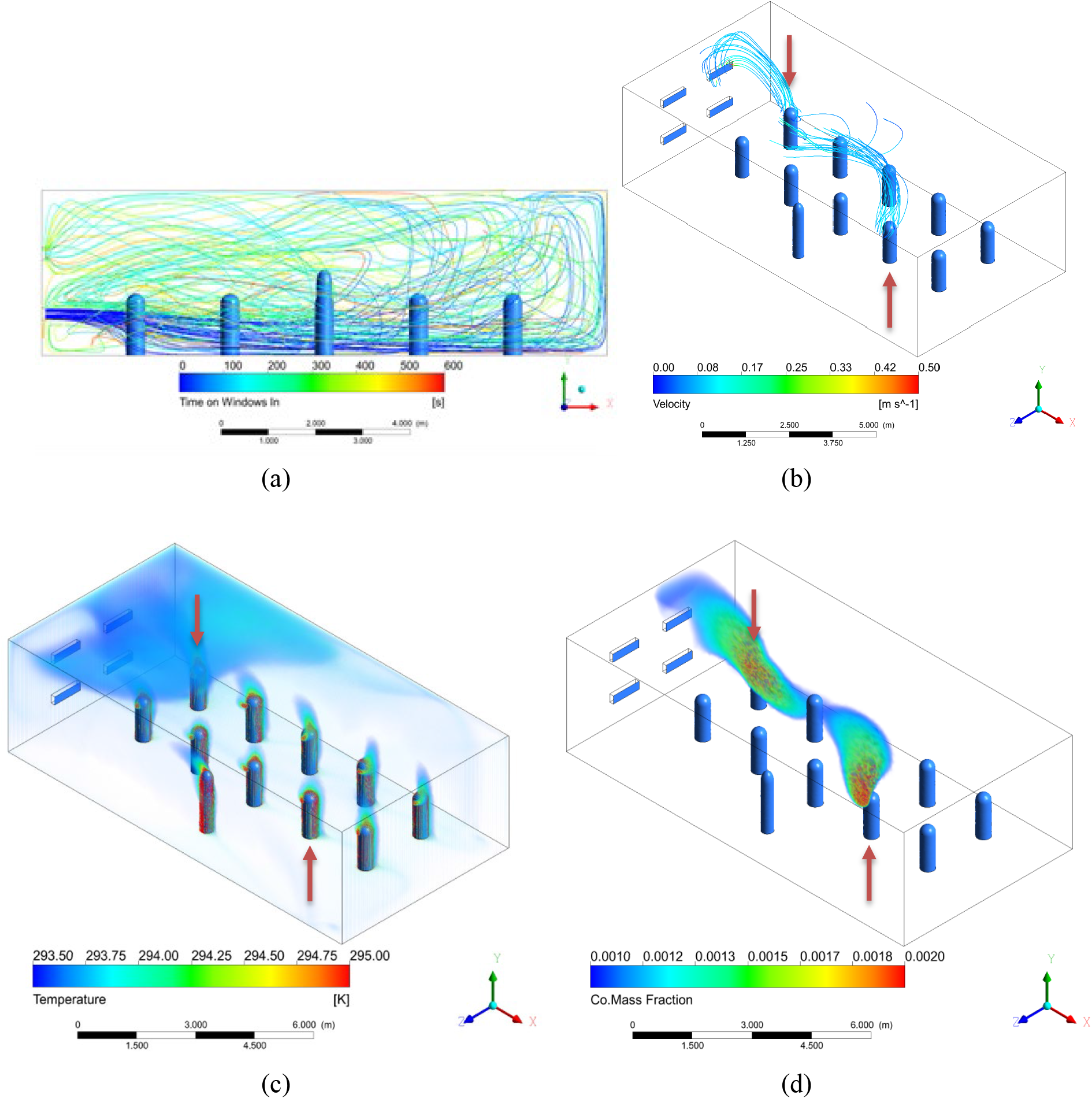
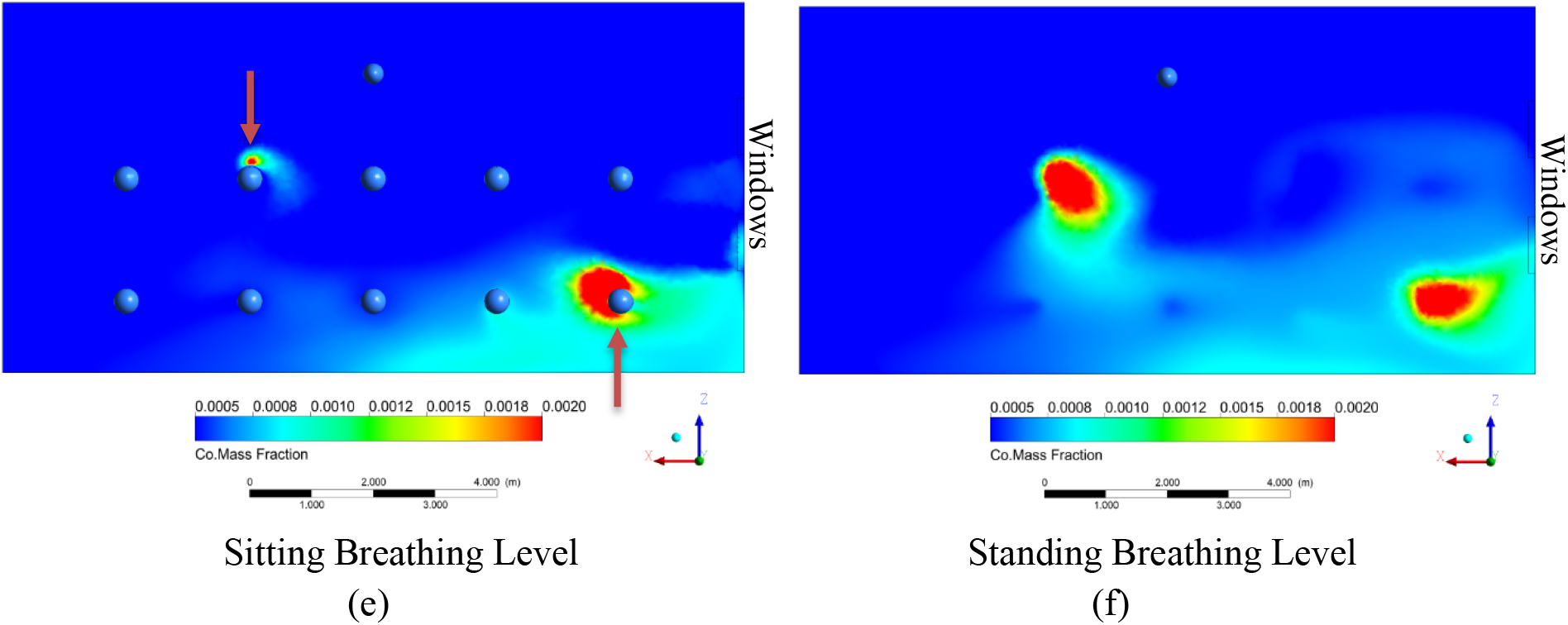
Results of Simulation B with double hung windows placed in between the rows of students. (a) Streamlines originating at bottom inlet windows from 0 to 600 seconds. (b) Streamlines originating around the heads of the two infected individuals from 0 to 60 seconds and colored according to velocity. (c) Volume rendering of temperature (K). (d) Volume rendering of CO mass fraction. The transparent limit is set at 2x well mixed, and the opaque scale at 4x well mixed. (e) Colored rendering of CO mass fraction right below mouth level of seated students. (f) Colored rendering of CO mass fraction right below mouth level of standing teacher. The lower limit is set at 1x well mixed, and the upper limit at 4x well mixed.

#### Simulation C: Baffles on Double Hung Windows Aligned with Rows of Students

Another possible way to avoid strong horizontal air currents interacting with exhaled air would be to place a baffle inside the windows that directs the air downwards. We simulated this by maintaining the same mass flow rate but directing the incoming air from the windows downwards. In Simulation C, the windows are again aligned with the rows of students as in Simulation A.

Compared with the results of Simulation A, where the air entered the room normal to the windows, the CO concentration at the breathing planes is lower for Simulation C, where the air from the windows is directed downwards. A cool layer of air near the floor seems to create a strong buoyancy effect so that most of the infected aerosols rise and are suspended above breathing level until they exit the room through the outlet windows. This starts to approach displacement ventilation although the air entering the room in this case has a high momentum causing recirculation and higher aerosol concentration at breathing level. These results suggest that baffles to direct air downwards could be an effective measure in classrooms with windows spanning an entire wall where the staggered alignment of simulation B is not possible.

#### Simulation D: Door Open and Bottom Window as Inlet

A strategy schools may employ to improve ventilation is to open the door along with the windows. In general, this would allow greater air flow into the room through the windows, since any positive pressure built up in the room can exit through the door. In Simulation D, we added an open door opposite of the windows. For this simulation, the windows are between the rows since we have established the risks of having them aligned with the students. The results are nearly identical regardless of if the windows are double hung since most of the air exits through the door. In this simulation, the upper window is closed.

Since the outlet is now on the opposite wall from the inlet and air can exit lower to the ground, a vertical temperature stratification occurs in the room with warm air accumulating above the top of the doorway, especially on the side of the room with the windows. Infected air rises with the human plumes and then becomes trapped in the warm layers as cool air continues to enter through the windows and creates buoyant effects. This situation is not ideal since the concentration is increased in the upper part of the room and the concentration of aerosols is elevated at the breathing level. If the windows were higher on the wall, the dangers of this situation may be reduced. This hypothesis is investigated in the next simulation.

#### Simulation E: Door Open and Top Window as Inlet

Simulation E, we again have a single open window with an open door on the opposite wall, however, the upper windows are now open, and the bottom windows are closed, creating a situation where the inlet is above breathing level.

The stratification seen in Simulation D was still prevalent in Simulation E, however the beginning of this warm layer was closer to the ceiling. The air flow entering from a higher elevated acted as a mixing force in the room, and more of the room was closer to the well-mixed CO mass fraction, as opposed to the large cloud of infected aerosols seen in Simulation D.

### HVAC Results

In the HVAC scenarios, the windows are sealed, and the room is ventilated from eight diffusers on the ceiling with a single return outlet near the top of the wall. The air enters along the ceiling from the diffusers with a speed of 0.5 m/s and temperature varying based on the simulation. The wall opposite the instructor is treated as a single glazed window, and several different outdoor temperatures are investigated. The results of the ceiling diffuser HVAC simulations are presented below. Table 2 shows different simulation conditions and their names. Table 3 contains their respective boundary conditions.

**Table 2:** Descriptions of simulation variations without operable windows

|  |  |
| --- | --- |
| Simulation F | Base Case Heating |
| Simulation G | Heating Higher Window Losses |
| Simulation H | Heating Highest Window Losses |
| Simulation I | Heating with Radiator |
| Simulation J | Cooling |
| Simulation K | Cooling More Window Heat |
| Simulation L | Higher Mouth Velocity Back Row |
| Simulation M | Higher Mouth Velocity Front Row |
| Simulation N | Adiabatic Window |

**Table 3:** Boundary conditions for simulations F-N, inclusive

| Simulation | F | G | H | I | J | K | L | M | N |
| --- | --- | --- | --- | --- | --- | --- | --- | --- | --- |
| **Inlet Temperature** | 301 K<br>(28 C) | 302 K<br>(29 C) | 303 K<br>(30 C) | 298 K<br>(25 C) | 288 K<br>(15 C) | 287 K<br>(14 C) | 301 K<br>(28 C) | 301 K<br>(28 C) | 292 K<br>(19 C) |
| **Window Heat Flux [W/m<sup>2</sup>]** | -50 | -75 | -100 | -75 | 35 | 70 | -50 | -50 | 0 |
| **Mouth Velocity [m/s]** | 0.35 | 0.35 | 0.35 | 0.35 | 0.35 | 0.35 | 1 | 1 | 0.35 |
| **Source Location** | Back Center | Back Center | Back Center | Back Center | Back Center | Back Center | Back Center | Front Right | Back Center |
| **Radiator Temp. – Air Temp. [K]** | Wall | Wall | Wall | 5 | Wall | Wall | Wall | Wall | Wall |

For the HVAC case, the total carbon monoxide injection was 0.0001544 kg/sec and the fresh air inlet was 0.343 kg/sec across the total interior air volume of 252 m^3^ yielding 4.4 air changes per hour. Based on this, the resulting well-mixed carbon monoxide concentration is 0.00045 total mass fraction of CO. The temperature of the inlet was adjusted to maintain comfortable temperature conditions in the room at steady state as the heat flux across the window changes. In these simulations, we assume an exhale velocity from the mouths of 0.35 m/s while maintaining the same mass flow rate as in the natural ventilation cases.

#### Simulation F: Moderate Heat Loss to a Window

This case represents the base heating case, with a moderate temperature gradient between the inside and outside. A negative heat flux of 50 W/m^2^ is present at the window along the back wall, corresponding to an outside air temperature roughly 10 K below the inside air temperature. Warm air enters the room from the ceiling diffusers at 301 K (28 C). Figure 10(a) below shows the presence of the thermal plumes around the individuals, which is key to the general airflow patterns in the room since there are no dominating horizontal air currents from windows. A similar visual, using CO mass fraction as the display variable, is shown in Figure 10(b). For the CO volume renders, the transparent boundary is set at 0.0005, which is 1.11 x the well-mixed concentration, and the red upper limit is set at 0.002, 4.44x well-mixed. From this visual we see that the effect of the thermal plumes has a significant effect on the spread of the exhaled gas, which rises around the infected individual, identified with an arrow.

**Figure 7:**
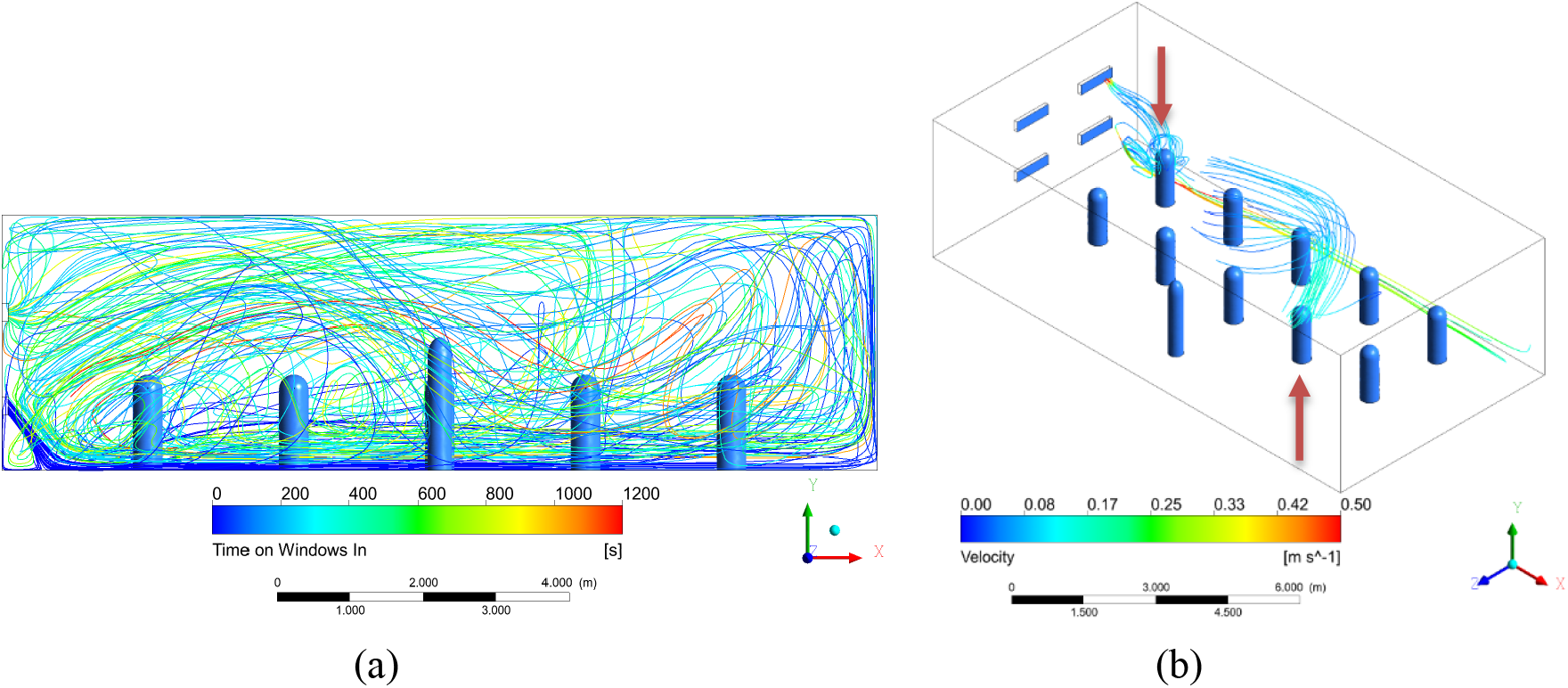
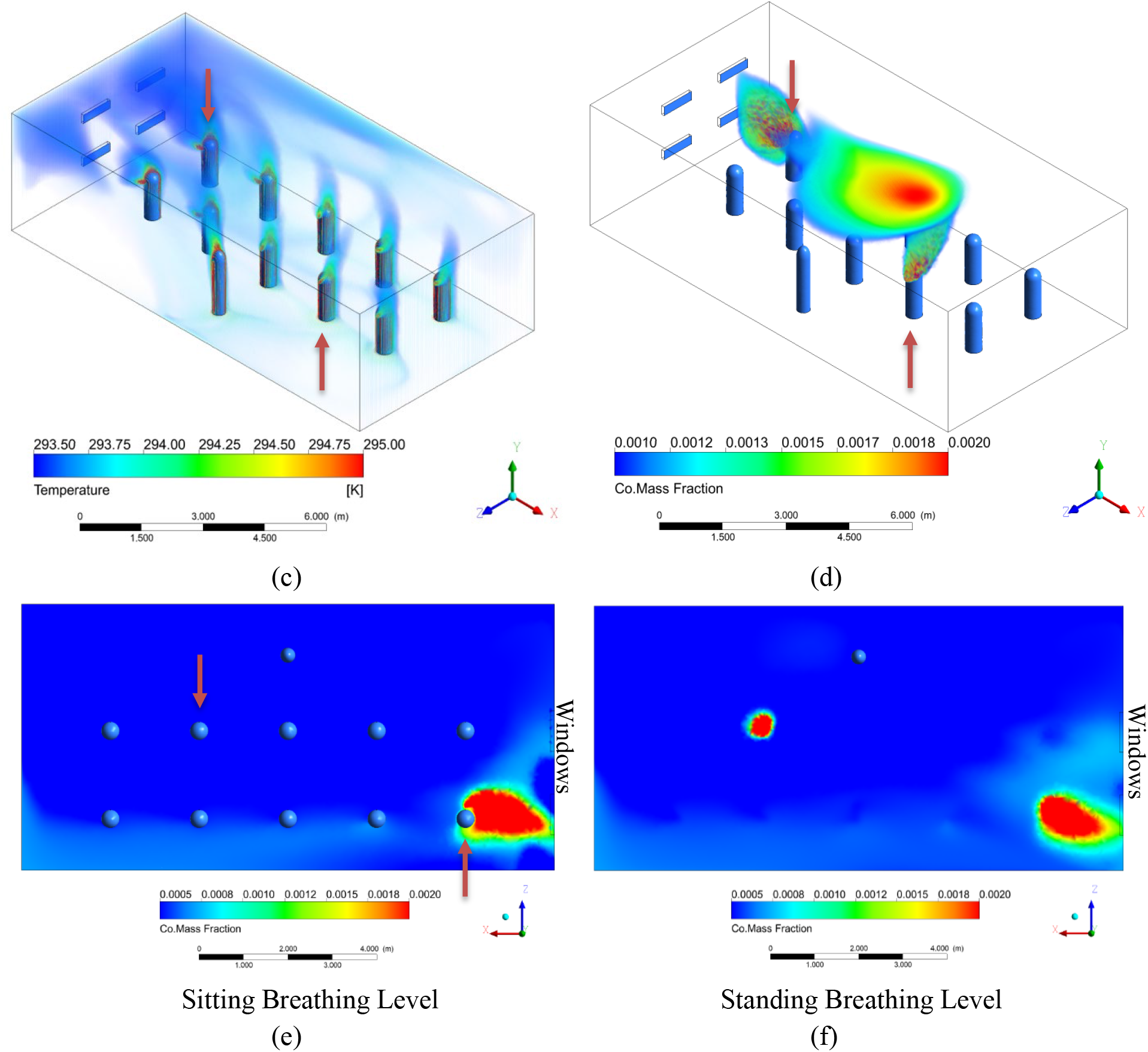
Results of Simulation C with double hung windows aligned with the rows of students and the air flow directed downwards to prevent strong horizontal drafts. (a) Streamlines originating at bottom inlet windows from 0 to 1200 seconds. (b) Streamlines originating around the heads of the two infected individuals from 0 to 60 seconds and colored according to velocity. (c) Volume rendering of temperature (K). (d) Volume rendering of CO mass fraction. The transparent limit is set at 2x well mixed, and the opaque scale at 4x well mixed. (e) Colored rendering of CO mass fraction right below mouth level of seated students. (f) Colored rendering of CO mass fraction right below mouth level of standing teacher. The lower limit is set at 1x well mixed, and the upper limit at 4x well mixed.

**Figure 8:**
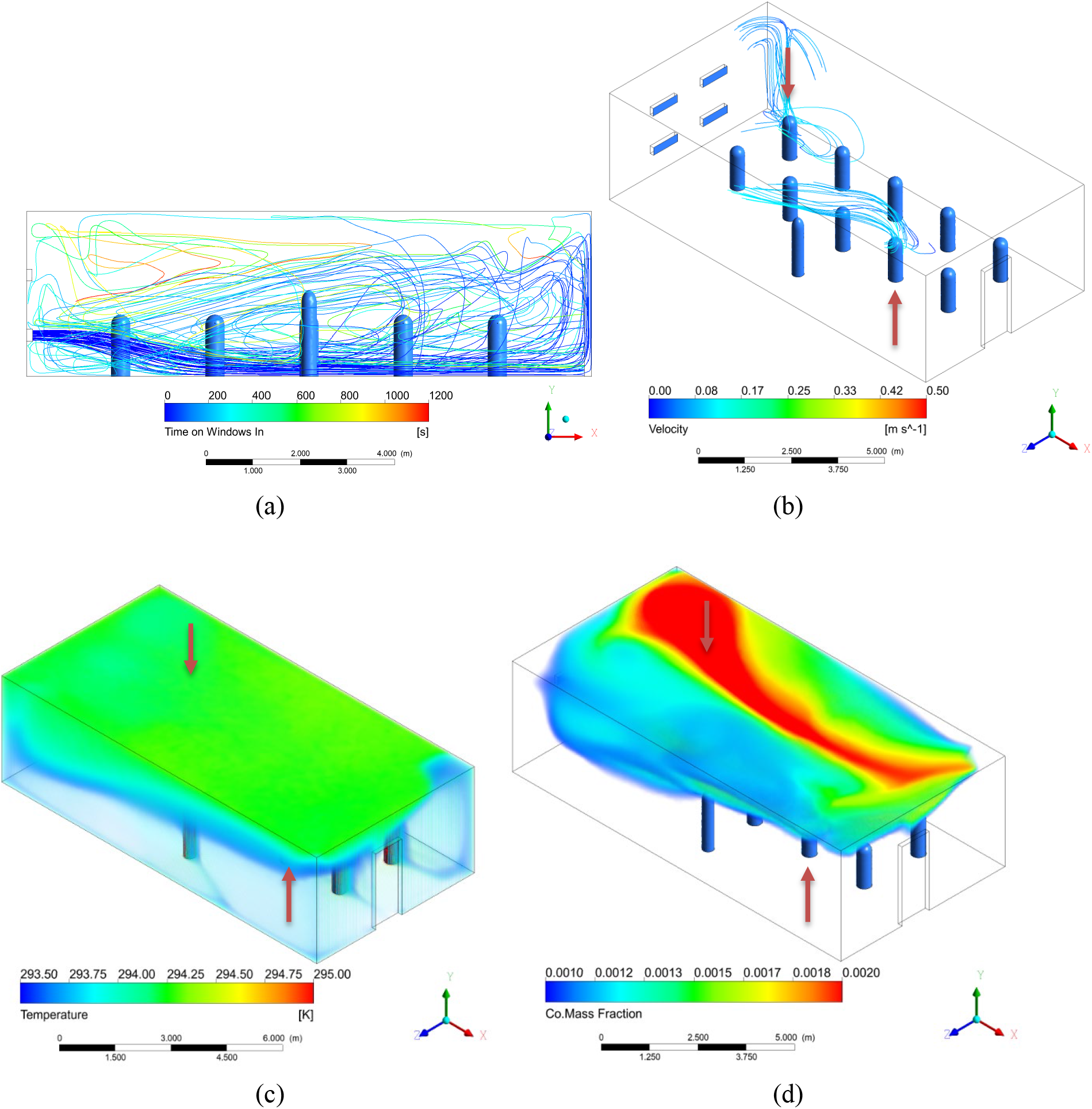
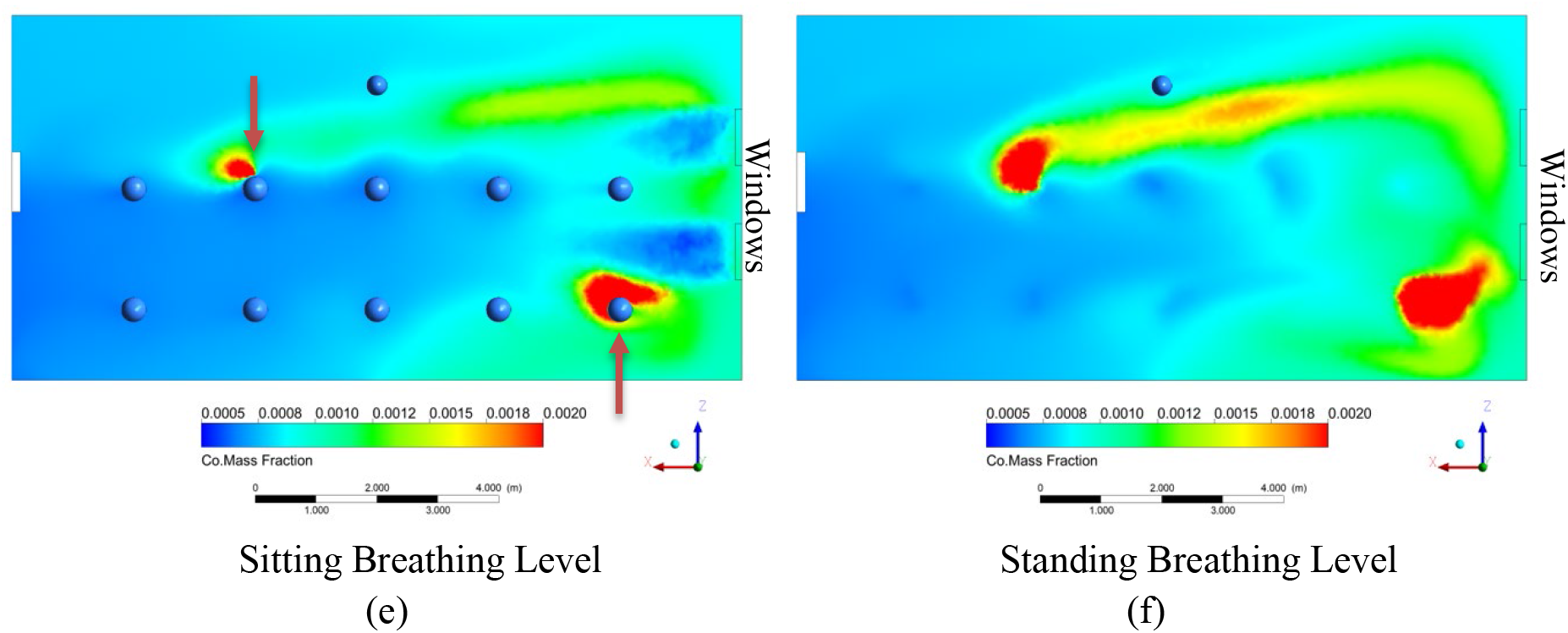
Results of Simulation D with a pair of windows between the rows of students and an open door on the opposite wall. Air flows in from the lower window. (a) Streamlines originating at bottom inlet windows from 0 to 1200 seconds. (b) Streamlines originating around the heads of the two infected individuals from 0 to 60 seconds and colored according to velocity. (c) Volume rendering of temperature (K). (d) Volume rendering of CO mass fraction. The transparent limit is set at 2x well mixed, and the opaque scale at 4x well mixed. (e) Colored rendering of CO mass fraction right below mouth level of seated students. (f) Colored rendering of CO mass fraction right below mouth level of standing teacher. The lower limit is set at 1x well mixed, and the upper limit at 4x well mixed.

**Figure 9:**
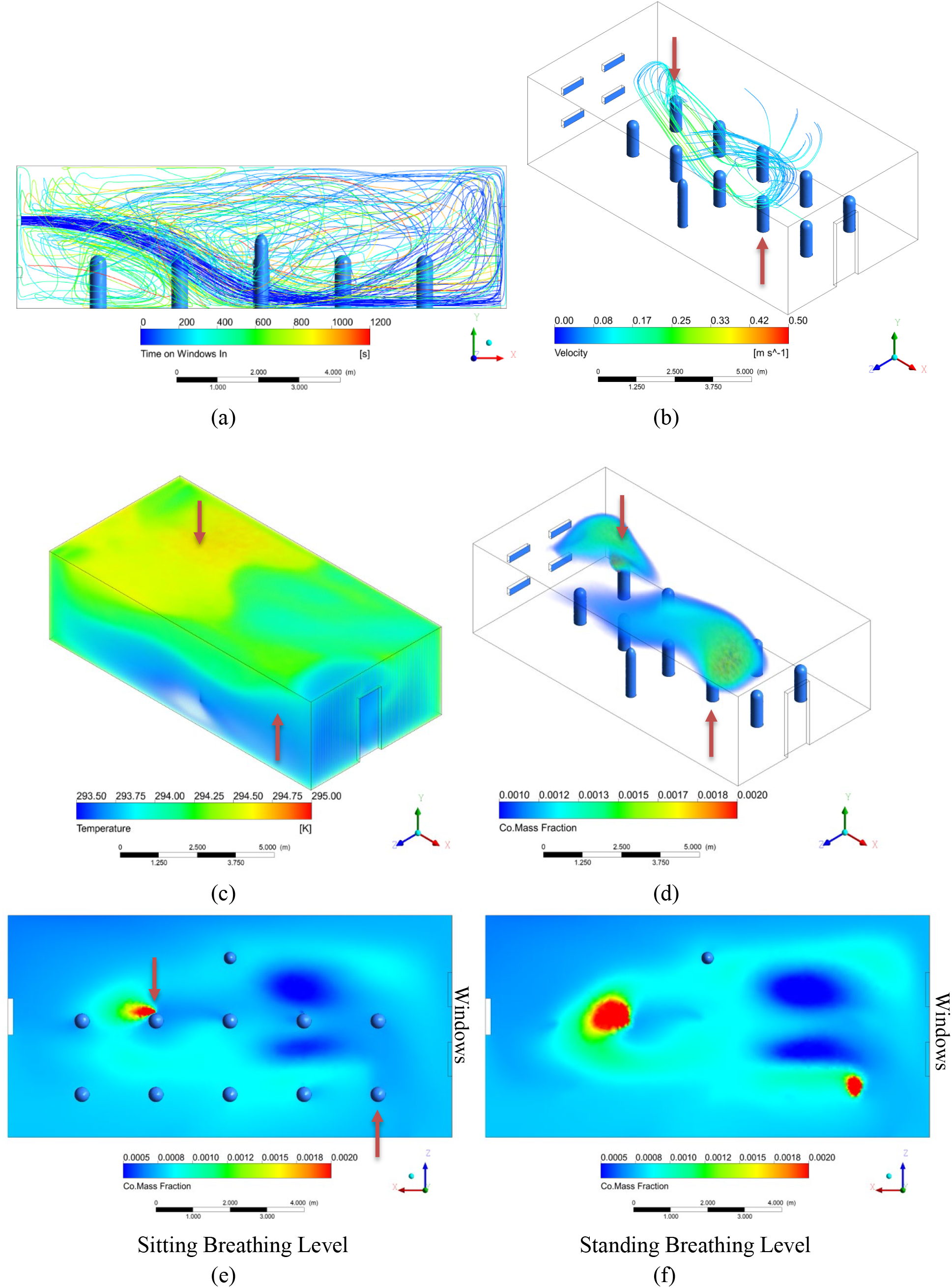
Results of Simulation E with a pair of windows between the rows of students and an open door on the opposite wall. Air flows in from the upper windows. (a) Streamlines originating at top inlet windows from 0 to 1200 seconds. (b) Streamlines originating around the heads of the two infected individuals from 0 to 60 seconds and colored according to velocity. (c) Volume rendering of temperature (K). (d) Volume rendering of CO mass fraction. The transparent limit is set at 2x well mixed, and the opaque scale at 4x well mixed. (e) Colored rendering of CO mass fraction right below mouth level of seated students. (f) Colored rendering of CO mass fraction right below mouth level of standing teacher. The lower limit is set at 1x well mixed, and the upper limit at 4x well mixed.

**Figure 10:**
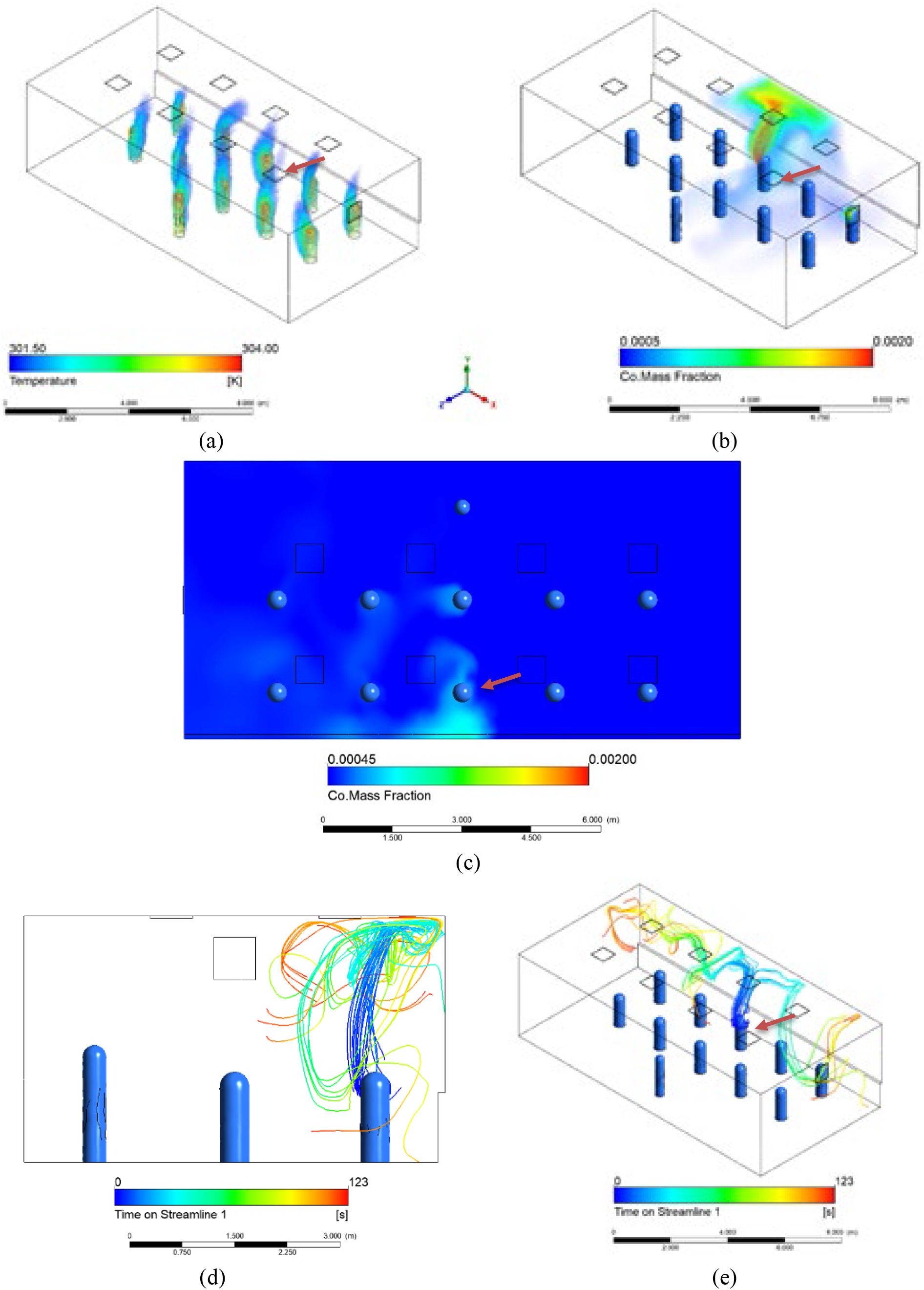
(a) Temperature Volume Render of Simulation F, clearly showing the thermal plumes in the room. (b) Volume Render of the CO mass fraction of Simulation F. (c) Horizontal Plane at breathing level, showing the CO mass fraction. (d, e) Steady state streamlines from infected individual for Simulation F.

We can see from these initial results that horizontal air flows are not as prevalent as in the natural ventilation cases, and the thermal plumes and cool air from the ceiling play a large role in the movement of air instead. The color scale on the streamlines in Figure 10(d, e) is representative of the time that has passed since that air passed by the infected individual. The streamlines clearly rise with the human plume before spreading throughout the room. The remaining sets of simulations will explore how changes in different variables impact the resulting steady-state conditions in the room.

#### Simulation G and H: Heating Case, Colder Windows

In Simulations G and H, the total heat flux at the window to the outside was increased to 75 W/m^2^ and 100 W/m^2^ to simulate larger temperature gradients across the single-glazed windows, analogous to a wintertime condition. The approximate outside temperatures for Simulations G and H are 10°C and 0°C respectively. From Figure 11, we can see that the increase of the temperature gradient resulted in much larger values of CO mass fraction throughout the room, indicating a more dangerous situation when an infected individual is seated next to a cool window with heating from above. The colder temperature at the window cools the adjacent air, which sinks to the floor from where it is picked up again by the plumes, carrying infected aerosols. The effect is magnified proportionally to the temperature gradient. This is shown in the streamline images as well as the CO volume renders in Figure 11, where the images on the left are Simulation G and the images on the right are Simulation H.

**Figure 11:**
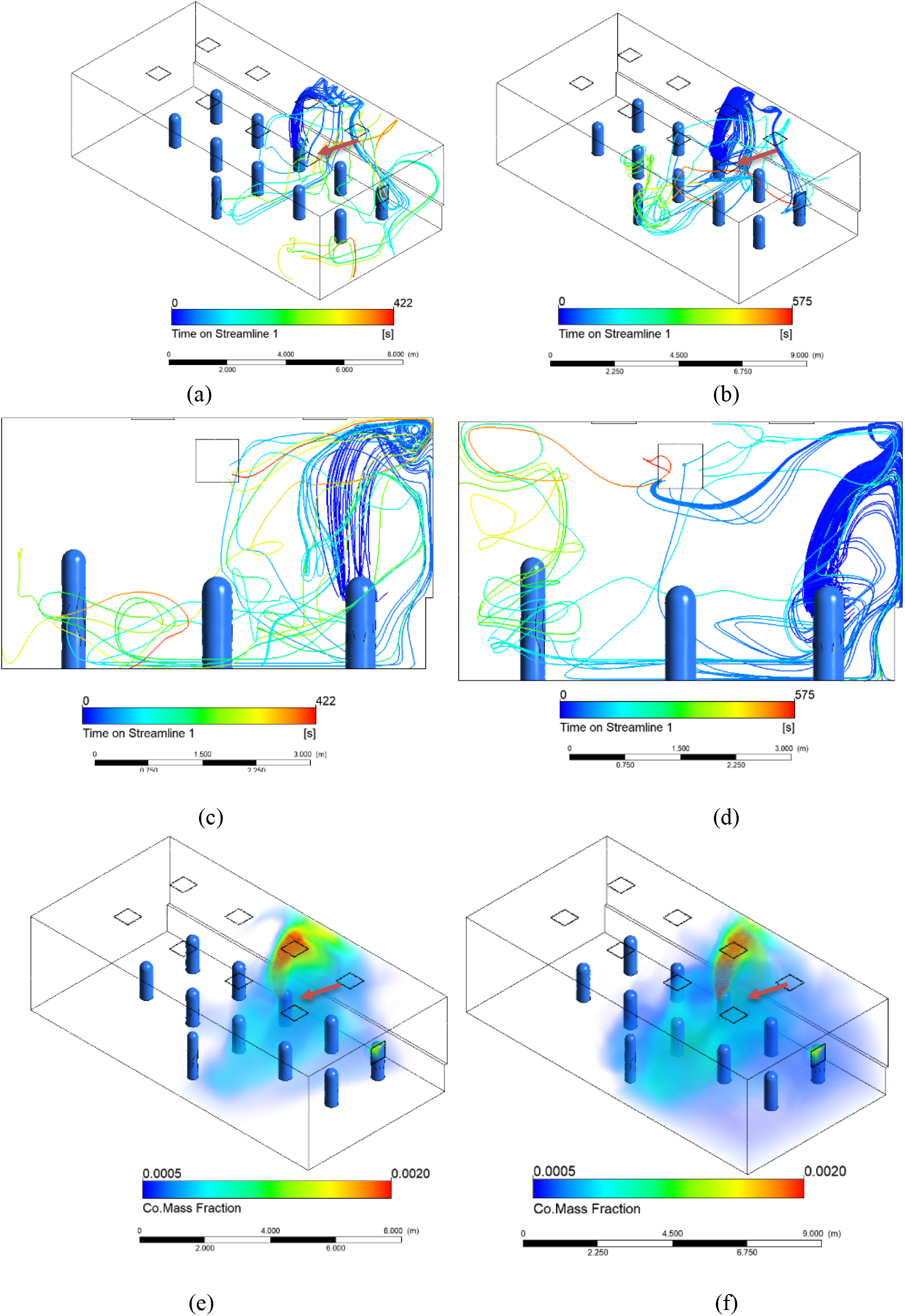
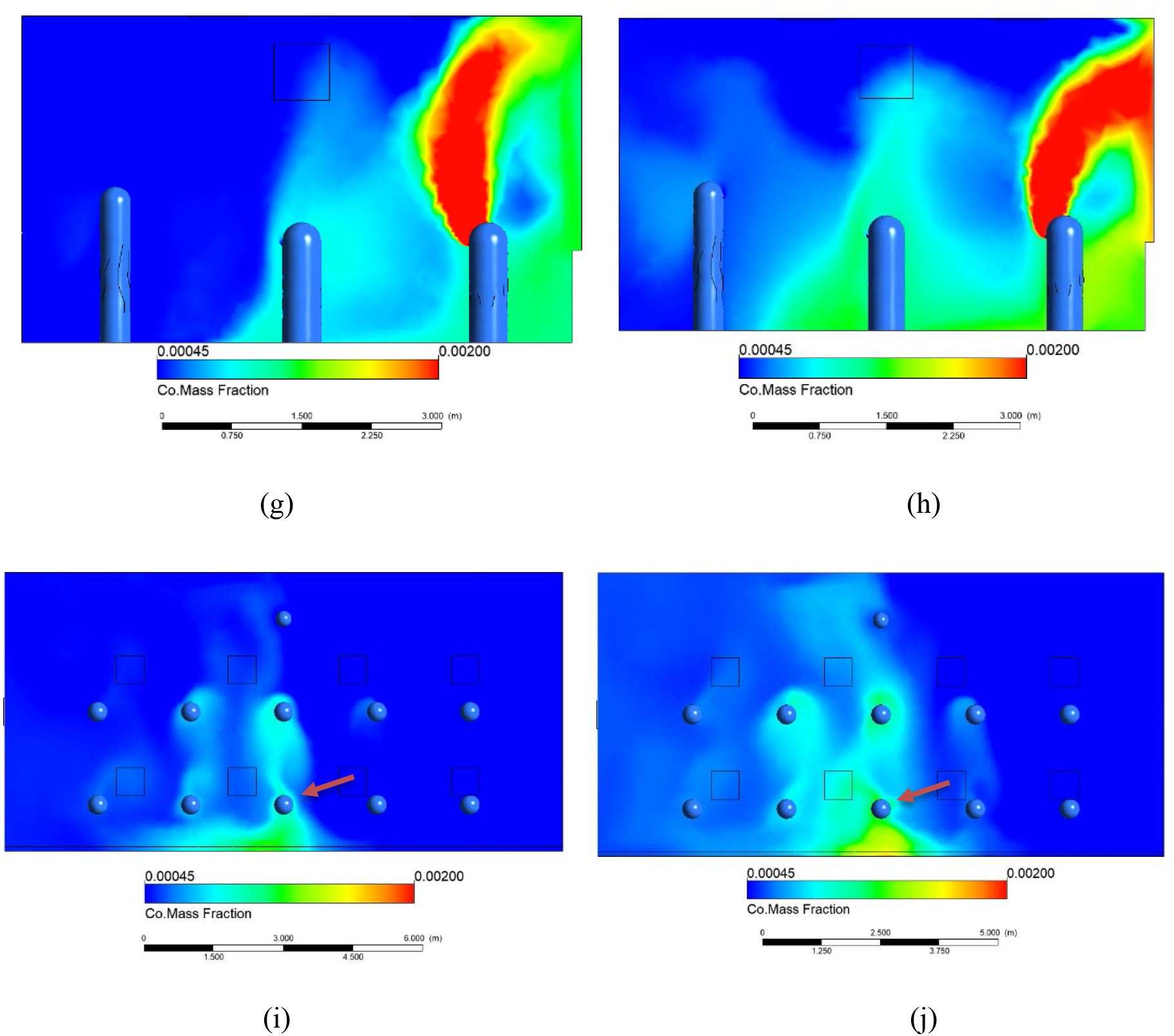
Simulations G and H are heating cases with outside temperatures 10° and 0°C respectively, represented by a negative heat flux across the back wall. Simulations G and H results. (a) Streamlines for Simulation G. (b) Streamlines for Simulation H. (c) Side view of streamlines for Simulation G. (d) Side view of streamlines for Simulation H. (e) Volume rendering of CO mass fraction for Simulation G. (f) Volume rendering of CO mass fraction for Simulation H. (g) Side view of CO mass fraction for Simulation G from plane taken through the center of the room. (h) Side view of CO mass fraction for Simulation H. (i) CO mass fraction at sitting breathing level for Simulation G. (j) CO mass fraction at sitting breathing level for Simulation H.

With the colder case, the higher aerosol concentration rises in the plume, is drawn down near the cold window, and then contacts the student in the next row. This effect of colder outside temperatures can be counteracted by implementing better thermal insulation at the window, such as double-glazing or using a radiator to heat the window surface. This last claim can be seen in the case for Simulation I in the box and whisker plot in Figure 14.

#### Simulations J and K: Cooling Cases, Warm Windows

In Simulation J the opposite season to Simulation F was considered, where the outside temperature is warmer than comfortable room temperature. From Figure 12, we can see that there is no large difference between the mild heating in Figure 10(c) and mild cooling case in Figure 12(e), but as soon as the net heat flux increases across the window, there are greater temperature gradients throughout the room, intensifying airflow patterns, and increasing airflow velocities. When the HVAC system compensates for a higher outside temperature, without adjusting the flow rate, the inlet temperature is decreased. This colder air, causes the air around the diffusers to sink down, including air which has risen in the infected plume. So as the inlet temperature decreases, the temperature gradient increases, causing more air to sink to the floor. This pattern is displayed in Figure 12, which shows the streamlines for the simulations J and K respectively.

**Figure 12:**
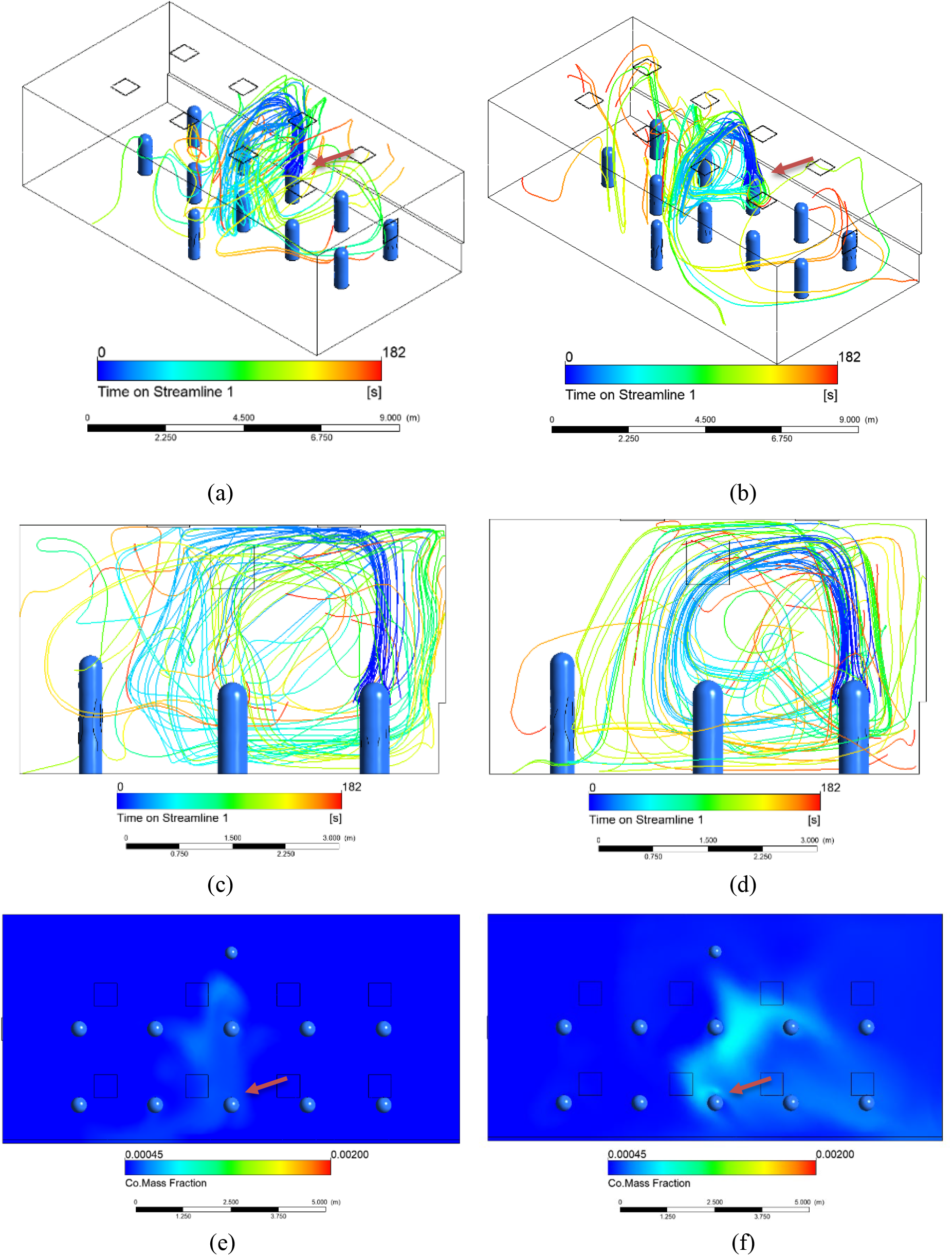
Simulations J and K are cooling cases with heat fluxes of 35 W/m^2^ and 70 W/m^2^ respectively, across the back wall. (a) Streamlines for Simulation J. (b) Streamlines for Simulation K. (c) Side view of streamlines for Simulation J. (d) Side view of streamlines for Simulation K. (e) CO mass fraction at sitting breathing level for Simulation J. (f) CO mass fraction at sitting breathing level for Simulation K.

#### Simulations L and M: Poorly Fitting Mask

In Simulations L and M, we investigate the effect that the velocity of exhaled air has on the CO mass fraction profile in the room. In these simulations, the velocity of the air exhaled by people was increased to 1 m/s, corresponding to people wearing a poorly fitting mask or not wearing a face mask at all. The infected individual in Simulation M is moved to the front row, to investigate dependence on location with respect to the window. In the CO mass fraction planes in Figure 13, the values are much higher than the other simulations, because the mouth velocity was increased without changing the geometry, the total CO volume introduced into the space increases proportionally. To provide adequate comparison between simulations, the scales for simulations L and M have been adjusted accordingly with respect to the well-mixed concentration. From the box and whisker plot in Figure 14, we can see that the normalized mass fraction values are higher for Simulations L and M than the lower velocity case in Simulation F. This further confirms our previous study on the effects of varied exhale speed in Figure 3. We can see that when the exhalation velocity is high enough, the exhaled air travels fast enough to escape the entraining effects of the thermal plume and spreads more by diffusion in the breathing plane.

**Figure 13:**
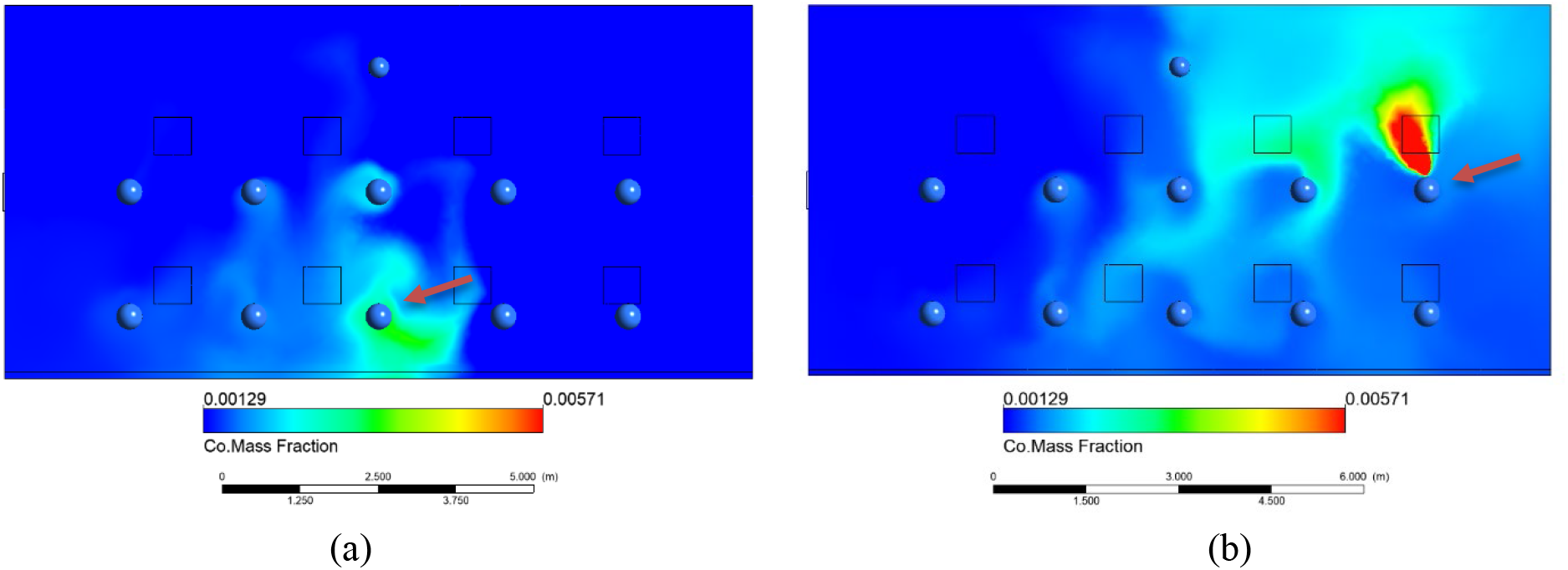
Simulations L and M for case of a higher breathing velocity than previous HVAC simulations. (a) CO mass fraction at sitting level for Simulation L with infected student in back row. (b) CO mass fraction at sitting level for Simulation M with infected student in front row on the far right. The upper limit of the color scale is 4.44x well mixed, and the lower limit is 1.11x well-mixed.

#### Simulation N: No Window, Adiabatic Room

In Simulation N, the thermal effects introduced by the window are removed to simulate a classroom with no windows. In this simulation, we see the CO tracer gas rises from the infected individual without much lateral motion. This further reinforces the theory of cold windows/walls creating worse conditions. From the comparison of all simulations shown below, we see the adiabatic wall case performing well. This suggests that better insulation on windows and walls leads to safer conditions inside the room.

#### Simulation Comparisons

The graphical results give a good qualitative picture of the aerosol dispersion. To obtain a quantitative comparison, a different method of comparison between situations was selected. By using CO as a tracer gas for exhaled air and monitoring the mass fraction throughout the space, we are able to gain some numeric basis for comparison. Samples of the average CO mass fraction, in a sphere of radius 0.33 m around the heads of all the non-infected individuals, were taken in each simulation. The results of these samples are shown in the box-and-whisker graph in Figure 14. The values are normalized with respect to the well-mixed CO mass fraction for each simulation. The distribution of CO mass fraction values can then be used as an indicator of infection risk for each of the situations. Cases A through E have ventilation through open windows while the rest of the cases use a standard HVAC ventilation design. The individuals with the highest and lowest concentration are shown as the extremes. Some cases resulted in an average local CO mass fraction value near the well-mixed value, whereas some averaged much higher. In some cases, the concentration reaches 2.5 times the well mixed value at certain locations.

**Figure 14:**
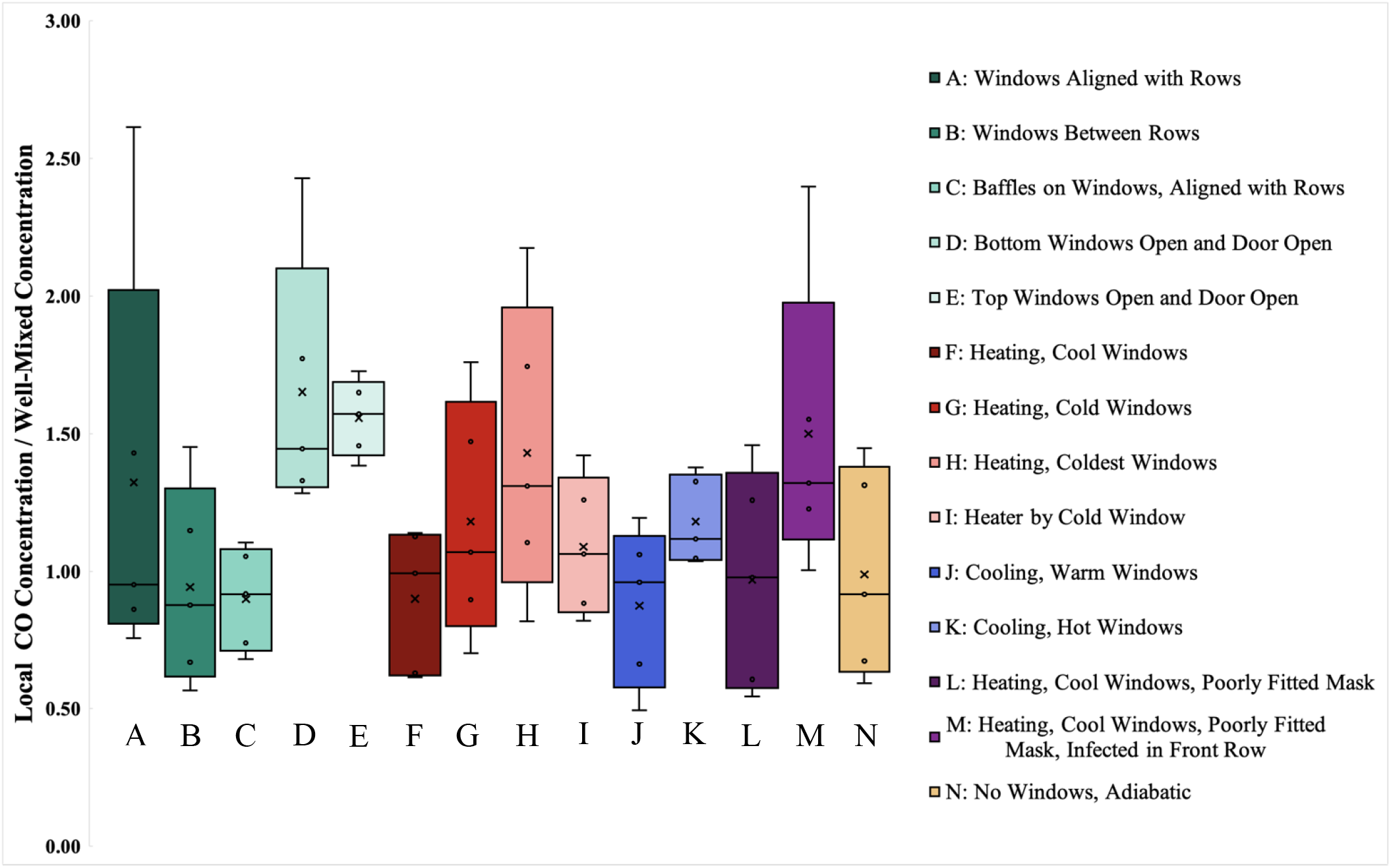
Box and whisker diagram of normalized CO mass fraction compared to well-mixed concentration at breathing plane for different simulations. ‘X’ represents the volume average concentration for the entirety of the room volume. The subset of values represented by the box and whisker plot represent individual spheres of 0.33 m radius surrounding the heat of the non-infected occupants within the space.

In some cases with open windows, the concentration reaches 2.5 times the well mixed value. The worst cases occur with lower windows in line with student seating and with exhaust through a modest door height. Directing the air immediately downward from the windows (mimicking displacement ventilation) gives the lowest level of exposure for all occupants. Opening the upper windows and the door results in a uniform exposure but at a higher average concentration.

For the HVAC cases with ceiling diffusers, concentration levels are highest when an infected individual is close to a very cold widow and when individuals wear a loosely fitted mask in a classroom with a cold window.

#### Flushing Simulation

A flushing simulation was completed on a transient solver. The same HVAC geometry with ceiling diffusers was modeled with adiabatic walls and an initial condition of the well mixed concentration at time zero. Occupants were intentionally not modeled during this simulation. This case is representative of a space being flushed out by the clean inlet air, overnight or between classes. If we assume large disturbances when the occupants leave the room, the well-mixed initial assumption is not far off. From the transient simulation, we found the average, maximum, and minimum volumetric mass fraction at 60 second intervals, and compared them to the corresponding theoretical, well-mixed values. There was a very close correlation between the two data sets. This suggests that a well-mixed assumption is an accurate representation of flushing an empty room.

### Energy implications and reduced occupancy

Based on this work, air change rate increases above the current recommendations are necessary to maintain exposure limits equivalent to the current well-mixed assumption for all occupants. Such an increase in air change rate will require a commensurate increase in energy requirements, to which local and zonal air filtration may be helpful in reducing contaminant levels near the source. In the simulations modeled above, the occupancy level has remained constant at eleven persons except for the flushing transient simulation. As shown in Figure 8(d), a wall-mounted CO_2_ sensor may also misrepresent the carbon dioxide levels in a space if the well-mixed assumption is invalid based on the airflow within same said space. The well-mixed assumption is particularly dangerous if the sensor location is used to control ventilation for the space itself.

As outside air is brought into a building for contaminant control, it is conditioned for both temperature and humidity limits, which requires energy to be spent in achieving those limits. A common control algorithm for the room HVAC system, demand control ventilation, maintains a prescribed CO_2_ level by varying the proportion of outside air in the ventilation airflow. The algorithm assumes a well-mixed steady state condition for the room, where the concentration of a pollutant, CO_2_ or aerosol, will be a function of the rate of pollutant mass generation and the mass flow rate of outside air.

With reduced occupancy, the total CO_2_ generation is reduced and the probabilistic chance of a non-infected individual encountering an infected individual is lower. However, with demand control ventilation and a reduced occupancy level, although the CO_2_ remains within prescribed limits, the resultant reduction of the outside air ventilation will result in a higher average concentration of aerosol in the room produced from an infected occupant (if present) than the same occupant in a room at full occupancy. Therefore, with reduced room occupancy, the chances of encountering an infected individual decreases. Yet if an individual is infected, the infection risk for other occupants increases with reduced room occupancy when demand control ventilation based on CO_2_ feedback is used. To address these scenarios, the safest operation requires a constant flow of outside air irrespective of occupancy level. To alleviate the energy impact of this strategy, an alternative approach may include a high-performance filtration system. Further work is needed to address this filtration question, as well as additional simulations of airflow to identify safe seating configurations with low occupancy and reduced airflow rates.

## Conclusions

COVID-19 is an aerosolized virus. With such small particle sizes, the established six-foot (∼1.8 m) spacing guidelines and heightened air change rates are an incomplete picture. The results from this paper help reinforce the concept that exhaled particles of modest velocity (such as a person with a mask or face shield) are entrained in the thermal plume of the body heat and rise toward the ceiling. However, higher horizontal velocities (such as a person without a mask or face shield) can escape the thermal plume and can linger within the breathing plane of others. These thermal plumes from people and heated objects continue to have an impact on air flow within the room, along with influences from cold or heated surfaces, mechanical HVAC systems, and outside air ventilation.

Two HVAC systems have been examined to show that, even with industry standard air change rates, the well-mixed assumption as applied to a conventional HVAC system made up of ceiling diffusers may underpredict the contaminant levels inhaled by the occupants by half. Even with the window open, the perceived safety of the well-mixed assumption is too conservative and underpredicts the potential exposure by 60%.

Aside from air change rate changes, other strategies can be employed to limit potential exposure of non-infected individuals. For situations where a cold outside surface (i.e. an uninsulated window) may exist, contaminant levels can be reduced by an order of 20% by the addition of a convective heater below the window, the use of curtains or blinds, or replacement of the windows to improve their insulative value. Care should be taken as a similar case is presented by the plastic enclosures used by many eateries where cold outside surfaces allow for recirculation of contaminants.

While open windows may give the impression of ventilation, they also create their own problems as the influx of air near the breathing plane carries contaminants horizontally from an infected person near the window to other occupants. Staggering seating arrangements and redirecting window air to the ground may remove this issue as it allows for the buoyant plumes to redevelop and bring contaminants out of the breathing plane.

Regarding flushing out contaminants in the absence of any occupants, the well-mixed assumption is valid for this application and a generalized air change model may be used.

Future work should include other HVAC systems such as displacement ventilation for improvements. For properly operated displacement ventilation systems with low speed floor inlets and ceiling exhaust, concentration at breathing levels should be below well mixed values. However, if occupants have loosely fitted masks, the performance could degrade if breathing exhaust escapes the thermal plumes around the individual. Cold windows could also degrade performance

Based on this work, air change rate increases above the current recommendations are necessary to maintain exposure limits equivalent to the current well-mixed assumption for all occupants. Such an increase in air change rate will require a commensurate increase in energy requirements, to which local and zonal air filtration may be helpful in reducing contaminant levels near the source. More work is necessary to determine the impact of increased ventilation on air flow patterns, the threshold for infection by aerosols, and to enhance the removal of aerosols through the design of the airflow within an occupied space. This work does not assess the impact of reducing occupancy levels further while occupied and represents an opportunity for further work to answer.

## Data Availability

CFD results are included within the manuscript. The workspace files are available upon request.

